# Informed decision making on the uptake of evidence-based smoking cessation assistance: A needs assessment among end users and experts to inform decision aid development

**DOI:** 10.1101/2021.04.09.21255012

**Authors:** Thomas Gültzow, Eline Suzanne Smit, Raesita Hudales, Carmen D. Dirksen, Ciska Hoving

## Abstract

**Introduction:** Evidence-based cessation assistance is known to increase cessation rates. Activating personal preferences as part of the decision for smoking cessation assistance tools could further improve tools’ effectiveness. Decision aids (DAs) help individuals to choose amongst the various options by taking these preferences into account and, therefore, could have a positive effect on cessation rates. To develop attractive and effective DAs, potential end users’ needs, and experts’ viewpoints should be considered during development processes. Therefore, the aim of this study was: (1) To explore smokers’ needs and viewpoints regarding a smoking cessation assistance DA, and (2) to obtain consensus among smoking cessation counsellors and scientific experts about the content and format of such a DA.

**Materials and methods:** Data was gathered via two approaches applied across three studies: (1) 20 semi-structured interviews with potential end users, (2) two three-round Delphi studies with 61 smoking cessation counsellors and 44 scientific experts. Data from the interviews and the first round of the Delphi studies were analysed qualitatively using the Framework method, while data from the second and third round of the Delphi studies were analysed quantitatively using medians and interquartile ranges.

**Results:** Potential end users reported to acquire information in different ways: Via own experiences, their social environment, and the media. Important characteristics to decide between tools also varied, however effectiveness and costs were commonly reported as the most important characteristics. The experts reached consensus on 38 (smoking cessation counsellors) and 40 (scientific experts) statements regarding important cessation assistance tools’ characteristics and their viewpoints on a smoking cessation assistance DA, e.g., that a tool should be appropriate for users’ level of addiction.

**Discussion and conclusion:** Some clear trends emerged among the potential end users (especially regarding important characteristics). Experts also reached consensus among a number of statements. However, there was some variation in the needs and wishes among the (different) stakeholders. The combination of these studies highlights that a ‘one size fits all’ approach is not desirable. In the development of DAs, this heterogeneity should be taken into account, e.g., by enabling users to customize a DA based on their personal preferences while safeguarding essential elements.

**Highlights:**

- Potential end users’ needs for a smoking cessation DA vary greatly
- However, tools’ effectiveness and costs were commonly named as important
- Customizable elements within a DA could be used to deal with this heterogeneity
- Conceptualizations (e.g., of effectiveness) may vary between stakeholders
- Information should be provided to end users in an easily understandable manner

## Introduction

Worldwide tobacco smoking continues to be the leading cause of preventable diseases and premature death (Samet, 2013). Evidence-based cessation assistance tools, e.g., nicotine replacement therapy (Stead et al., 2012) or smoking cessation counselling (Matkin et al., 2019), are known to increase successful smoking cessation chances. Zhu et al. (2000) reported that cessation assistance tools double the chances of attaining a smoke-free status. However, evidence-based cessation assistance tools are currently underused (Cokkinides et al., 2005). Increasing the uptake of said tools would therefore likely result in more individuals achieving smoking abstinence, which can lead to improved population health, as well as a decrease in healthcare costs (Kahende et al., 2009; Ruger & Lazar, 2012).

One commonly described barrier to using evidence-based cessation assistance tools are incorrect beliefs regarding their safety and efficacy (Bansal et al., 2004), while adequate knowledge is one of the prerequisites for making informed decisions (Bekker et al., 1999; van den Berg et al., 2006). Conversely, this means that increasing individuals’ knowledge about evidence-based cessation assistance tools could lead to more informed decisions and potentially result in more individuals using said assistance during a quit attempt. However, even if individuals are knowledgeable and do choose to seek additional help in the form of cessation assistance tools, the variety of effective tools (e.g., Cahill et al., 2013; Rice et al., 2013; Stead et al., 2013; Taylor et al., 2017) means that they still have to make a decision between the different effective tools available. As making choices based on personal preferences has been shown to positively influence the effectiveness of clinical treatments (Brazier et al., 2009), incorporating personal preferences when deciding on which cessation assistance support tool to use could be beneficial for smoking cessation as well. In addition to providing information to expand knowledge, it could, therefore, be useful to help individuals to identify what is important to them personally (i.e., help them to clarify their values) and support them in choosing a cessation aid that fits their personal preferences – another prerequisite for making informed decisions (Bekker et al., 1999; van den Berg et al., 2006).

Decision aids (DAs) are interventions specifically meant to provide unbiased information in order to increase users’ knowledge, and to support these users in choosing a healthcare option that best reflects their personal values and preferences (Stacey et al., 2017) – in other words: DAs support informed decision making. DAs have predominantly been developed to support people in making decisions about medical treatments and screening programs (Stacey et al., 2017), but are also increasingly used to help smokers make informed decisions regarding preventive health-related behaviours, such as smoking cessation (Gültzow et al., 2021; Moyo et al., 2018). In various studies it has been shown that DAs can have positive effects on smoking cessation outcomes (e.g., BinDhim et al., 2018; Cupertino et al., 2010; Willemsen et al., 2006), however, as far as we are aware, in all of those studies assessments of needs and viewpoints of potential end users in initial phases of DA development were not reported. This is in contrast to the criteria of the International Patient Decision Aid Standards (IPDAS) Collaboration (Durand et al., 2015) that explicitly recommend this assessment in the developmental process of any DA. This is unfortunate, as including potential end users early on in the development process may lead to increased end user acceptance, and improved long-term implementation and effectiveness of DAs (Coulter et al., 2013; Hooiveld et al., 2018; Vaisson et al., 2021). Within the IPDAS development guidelines, the assessment of clinician’s viewpoints is also advocated (Coulter et al., 2013) which mainly is a reflection of the fact that DAs are traditionally employed in clinical settings (Stacey et al., 2017; Vaisson et al., 2021). Given that decisions on health promotion and public health issues (such as smoking cessation (Borland et al., 2012)) are commonly made without the involvement of clinicians, the question is whether different or additional experts (such as scientific experts) should play a role in the development of DAs that serve a health promotion or public health goal. There are currently no guidelines on how to develop such DAs and, consequently, there is no consensus on which experts should be involved in the development process. Therefore, we chose to expand the commonly accepted approach to include clinicians (in our specific case, smoking cessation counsellors) with the inclusion of additional experts, i.e., scientific experts in the field of smoking cessation.

Therefore, the aim of this study was to explore the needs and viewpoints regarding the development of a DA to support informed decision making within the context of smoking cessation from the perspective of three main stakeholders: (1) Potential end users (i.e., individuals motivated to quit smoking), (2) smoking cessation counsellors and (3) scientific experts. In order to optimally explore the needs and viewpoints of the different stakeholder groups, two different approaches were applied in three studies: (1) In-depth, semi-structured interviews were conducted to identify the smokers’ needs and wishes in relation to the decision-making process on how to quit smoking and the content and format of an online DA that aims to support them in this process; (2) two three-round Delphi studies among (1) smoking cessation counsellors and (2) scientific experts, to gather their viewpoints and experiences regarding smoking cessation decision making, and to obtain consensus on the required content of an online smoking cessation DA. The result from these studies were directly applied by the research team in order to develop a smoking cessation DA (Gültzow, Smit, Hudales, Knapen, et al., 2020). However, the results of the three studies described in this article can also be used to inform the development of other DAs, related to smoking cessation or other preventive health-related behaviours, and offer new insights into the decision-making process that smokers go through when they decide how they want to quit smoking.

## Material and methods

### Study design

As indicated in the introduction, this study was part of a larger project with the ultimate goal of developing an online DA to support people in making an informed decision about the use of cessation assistance tools (Gültzow, Smit, Hudales, Knapen, et al., 2020). This larger project is funded by the Dutch Cancer Society, UM2015-7744. Evaluation of this project by the Medical Ethics Committee METC Z (16-N-227) revealed that this project did not require medical ethics approval under the rules of the Medical Research Involving Human Subjects Act (WMO). All materials that participants received, interview guides, questionnaires and SPSS syntaxes can be found on the Open Science Framework (OSF) (https://osf.io/6dg7r/?view_only=98e511e58fcd4870a583c2071430e526). For the interview study, we only produced materials in Dutch, however, to facilitate understanding we provided approximate translations.

### In-depth semi-structured interviews with individuals motivated to quit smoking

#### Participants and procedure

For the interview study, we recruited among members from a national research panel (Flycatcher Internet Research, 2018) that participated in an earlier study about smoking cessation assistance decision making (Gültzow, Smit, Hudales, Dirksen, et al., 2020). We employed purposeful sampling to invite a heterogenic sample using the following four characteristics: (1) Intention to use the proposed online DA (Ajzen, 1991), (2) decision-making style as measured by the General Decision Making Style measurement (Scott & Bruce, 1995), (3) geographical location and (4) reported gender identity (i.e., man, woman and non-binary). Interviews were conducted until data saturation was reached (Mason, 2010), i.e., when three consecutive interviews did not generate new knowledge related to the research objectives.

All interviews were conducted by telephone and recorded, for which participants gave their consent after a brief explanation about the study and the procedure. Prior to the actual interview, participants received: (1) A description of the different decision-making styles based on the aforementioned earlier study (see Gültzow, Smit, Hudales, Dirksen, et al. (2020) for more information), (2) information about which cessation assistance tools are evidence-based (e.g., behavioural support) and which are not (e.g., acupuncture), and (3) DA mock-up screenshots to give an indication of what an online DA to support people in making an informed decision about the use of cessation assistance could look like. Participants received a € 30 ($ 32,63) gift card for their participation.

#### Interview guide

A semi-structured interview guide was developed which was used to gather information for the larger project (Gültzow, Smit, Hudales, Knapen, et al., 2020) and this study, and therefore also included questions that were beyond the scope and objectives of this study (see the entire interview guide on the OSF, https://osf.io/6dg7r/?view_only=98e511e58fcd4870a583c2071430e526). The relevant questions for this study were related to: (1) Knowledge, (2) values related to cessation assistance tools, (3) the proposed DA (e.g., personal needs regarding functional aspects, device compatibility, usage duration and the layout of the DA). Two nearly identical interview guides were developed; one for participants that were still smoking and one for smokers who recently quit (e.g., questions were posed about the last quit attempt instead of future attempts).

#### Data analysis

Data was analysed using the Framework Method (Gale et al., 2013) using NVivo 12 (QSR International, 2018). After interviews were transcribed verbatim by RH, TG and RH created a codebook in an iterative process. TG and RH used the final version to code all interviews (TG) and 10% (n=2, RH) respectively, resulting in a high intercoder reliability of 0.81 (Cohen’s Kappa) with 99.5% of agreement. After intercoder reliability was ensured, data was interpreted by making use of a framework matrix to look for patterns in relation to knowledge, values, and the proposed online DA. In a framework matrix, qualitative data is summarized and analysed. This enables easy comparison between the participants to identify patterns (Gale et al., 2013). Additionally, we analysed if patterns could be observed based on gender identity, age and level of education. Sample characteristics were examined using SPSS (IBM Corp, 2017).

### Three-round Delphi studies among scientific experts and smoking cessation counsellors

#### Participants and procedure

For the Delphi studies, we used different recruitment approaches for the two expert groups. Smoking cessation counsellors were included if they offered counselling services to smoking citizens. We mainly drew from our professional networks, relevant Dutch organizations, newsletters and we employed snowball recruitment (i.e., participants were asked to name up to three additional experts).

Scientific experts’ recruitment occurred through a literature search, mass media channels (e.g., newsletters and social media) and (again) snowballing (i.e., participants were asked to name up to three additional experts). We used database searches in PubMed to identify relevant papers using the following search terms: “(smoking cessation OR tobacco cessation) AND (intervention OR program*)”. The search was limited to the past five years and to journal articles written in English or Dutch. First, second and last authors of papers relevant to the development, testing and/or implementation of smoking cessation interventions were invited by email to participate in our study. In order to ensure that participants had enough relevant expertise five inclusion criteria (based on Davis et al. (2004)) were used: Participants had to (1) have published at least two journal articles/book chapters or one book on smoking (cessation) and/or smoking cessation interventions within the last 5 years, (2) have been awarded a grant within the last five years in the area of smoking (cessation) and/or smoking cessation interventions, (3) have been involved in at least one development process of a smoking cessation intervention within the last 5 years, (4) have at least five years of experience in the field of smoking (cessation) and/or smoking cessation interventions. Participants had to fulfil at least two criteria to be included in the first round (as the answers from the first round form the basis for the following rounds – see *The questionnaires*), and at least one to be included in the other rounds.

#### The questionnaires

The questionnaires of all rounds consisted of questions pertaining to (1) the professional function and background of participants, (2) the inclusion criteria (see *Participants and procedure*), and (3) participants’ email addresses. Starting with round two, experts were also asked if they have been involved in the development of at least one DA. Additionally, five open-ended questions were posed in the first round *(Table 1)* and one in the second and third round (“If you have any additions or comments, please feel free to add them here.”).

**Table 1.**
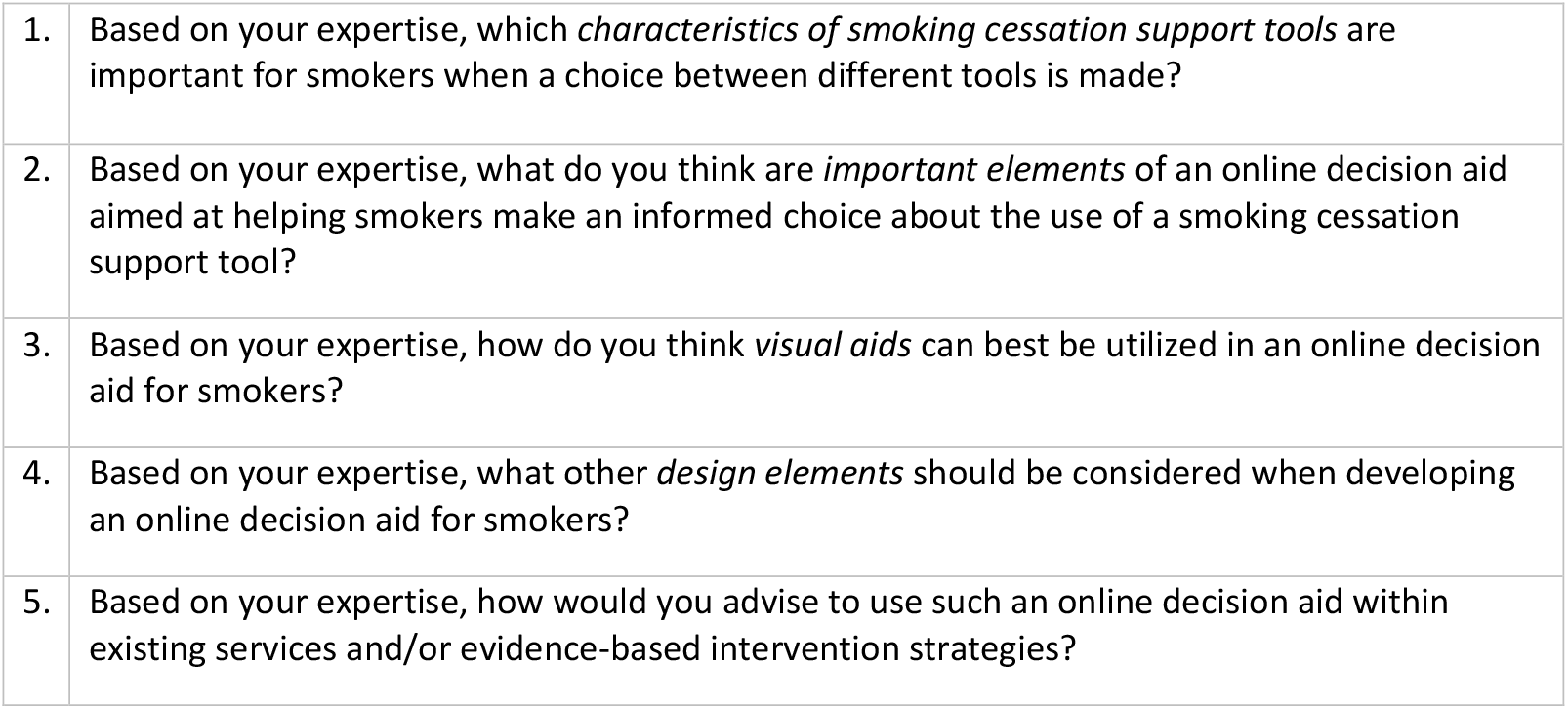
Questions first round.

|  |  |
| --- | --- |
| 1. | Based on your expertise, which *characteristics of smoking cessation support tools* are important for smokers when a choice between different tools is made? |
| 2. | Based on your expertise, what do you think are *important elements* of an online decision aid aimed at helping smokers make an informed choice about the use of a smoking cessation support tool? |
| 3. | Based on your expertise, how do you think *visual aids* can best be utilized in an online decision aid for smokers? |
| 4. | Based on your expertise, what other *design elements* should be considered when developing an online decision aid for smokers? |
| 5. | Based on your expertise, how would you advise to use such an online decision aid within existing services and/or evidence-based intervention strategies? |

The questions of the first round were deliberately designed to align with the questions asked to the end users to facilitate comparison between the different groups. The questionnaire of the second round included 75 statements and was based on the answers of the first round (and other literature for two statements to supplement the experts’ answers) regarding (1) Cessation assistance tools’ characteristics, (2) online DA functions, (3) visual aids, (4) embedment of the DA in the current clinical context. After answering statements regarding cessation assistance tool’s characteristics, participants were shown DA mock-up screenshots to give them an indication of what an online DA could look like based on input from the first round to provide them with context before answering the rest of the statements. All statements were scored on a 7-point Likert scale (see *Table 4* for more information).

**Table 4.**
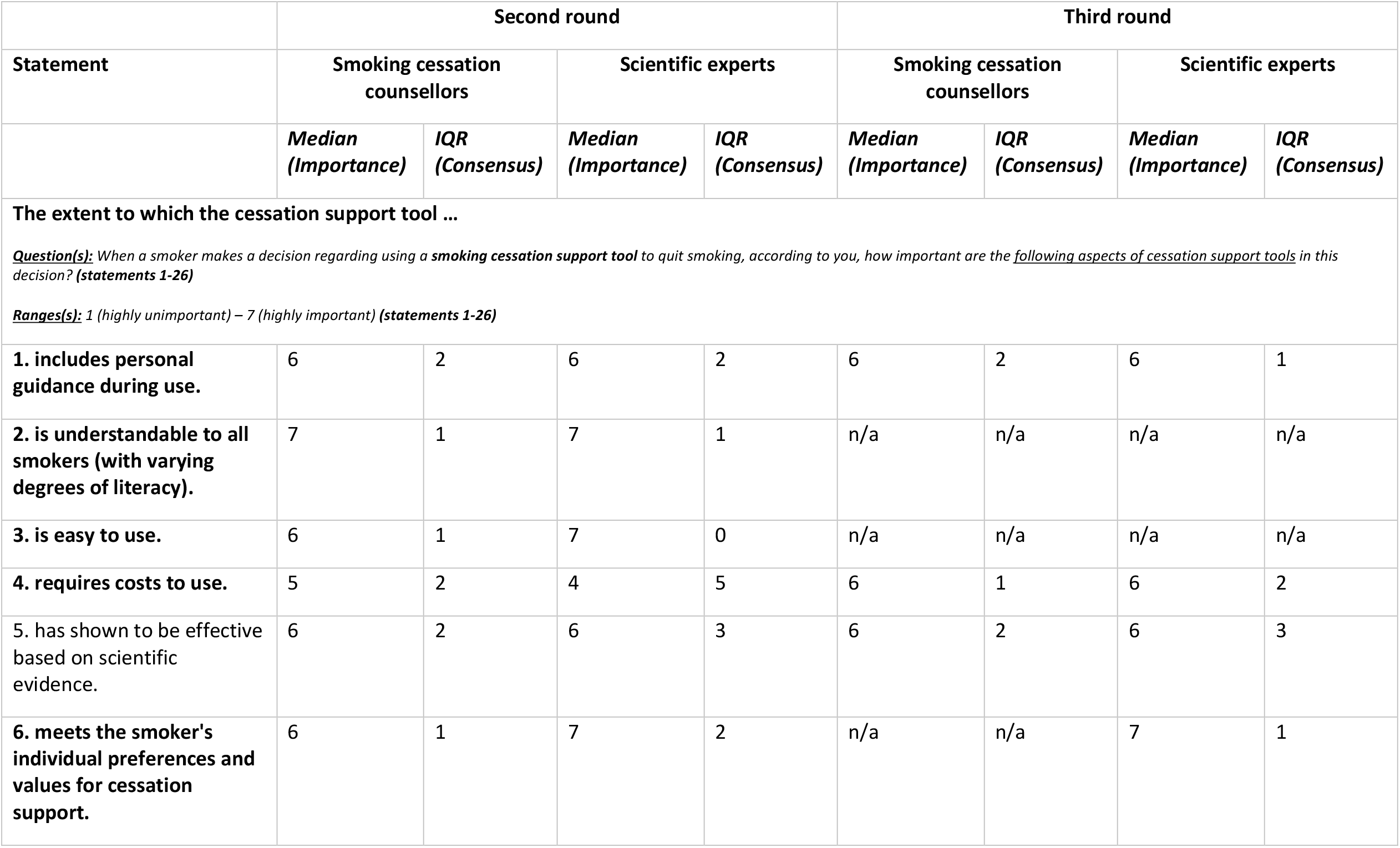
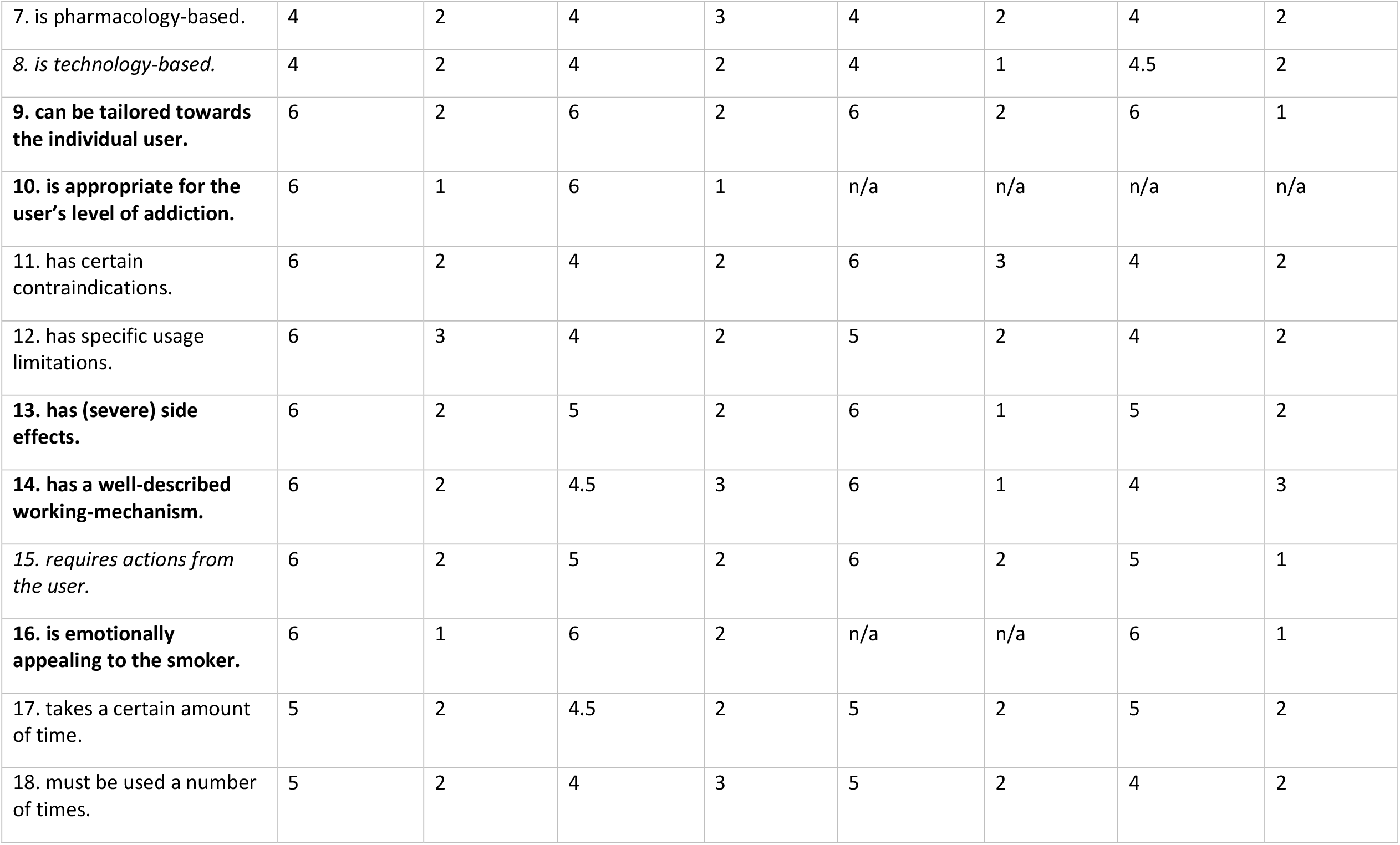
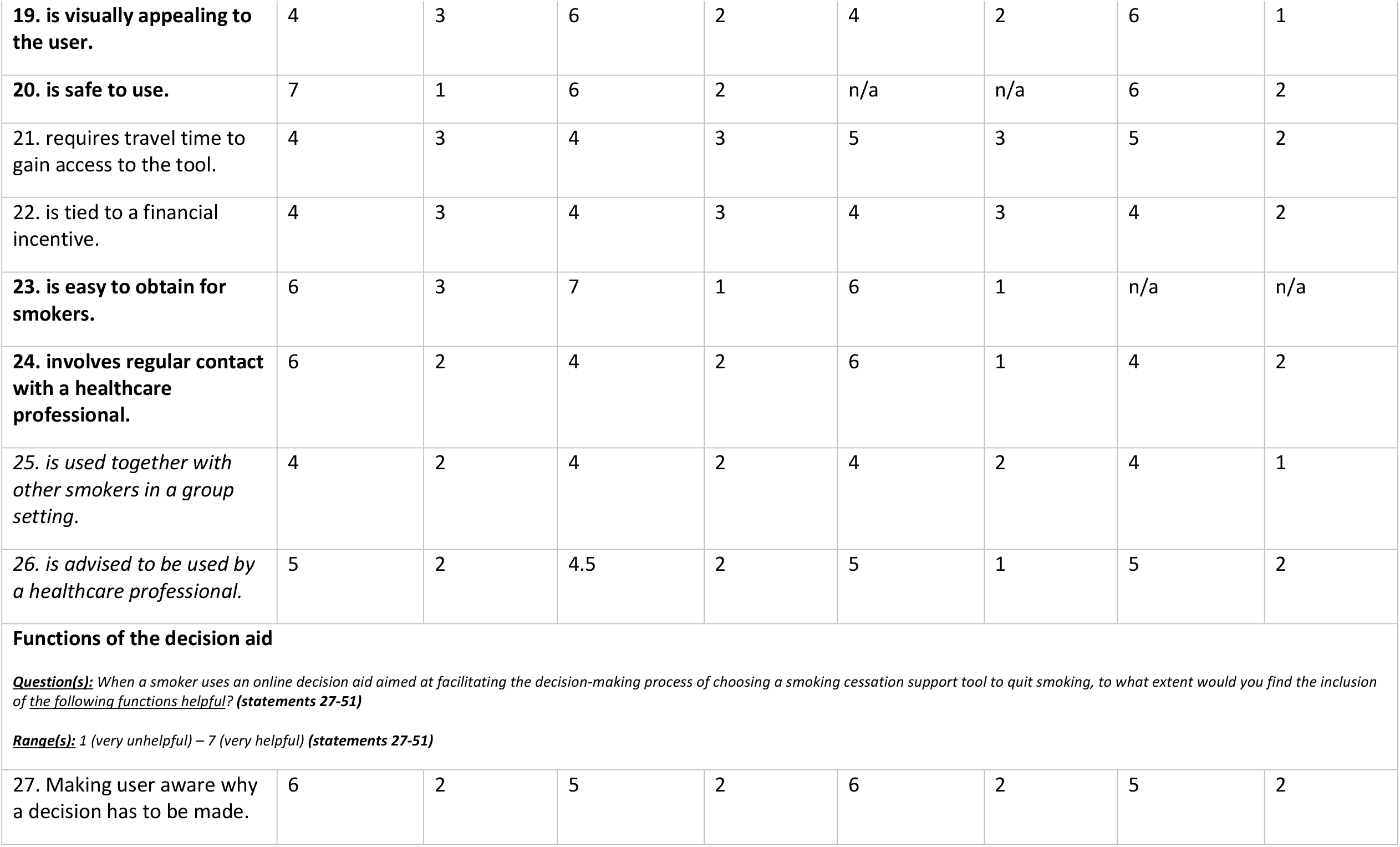
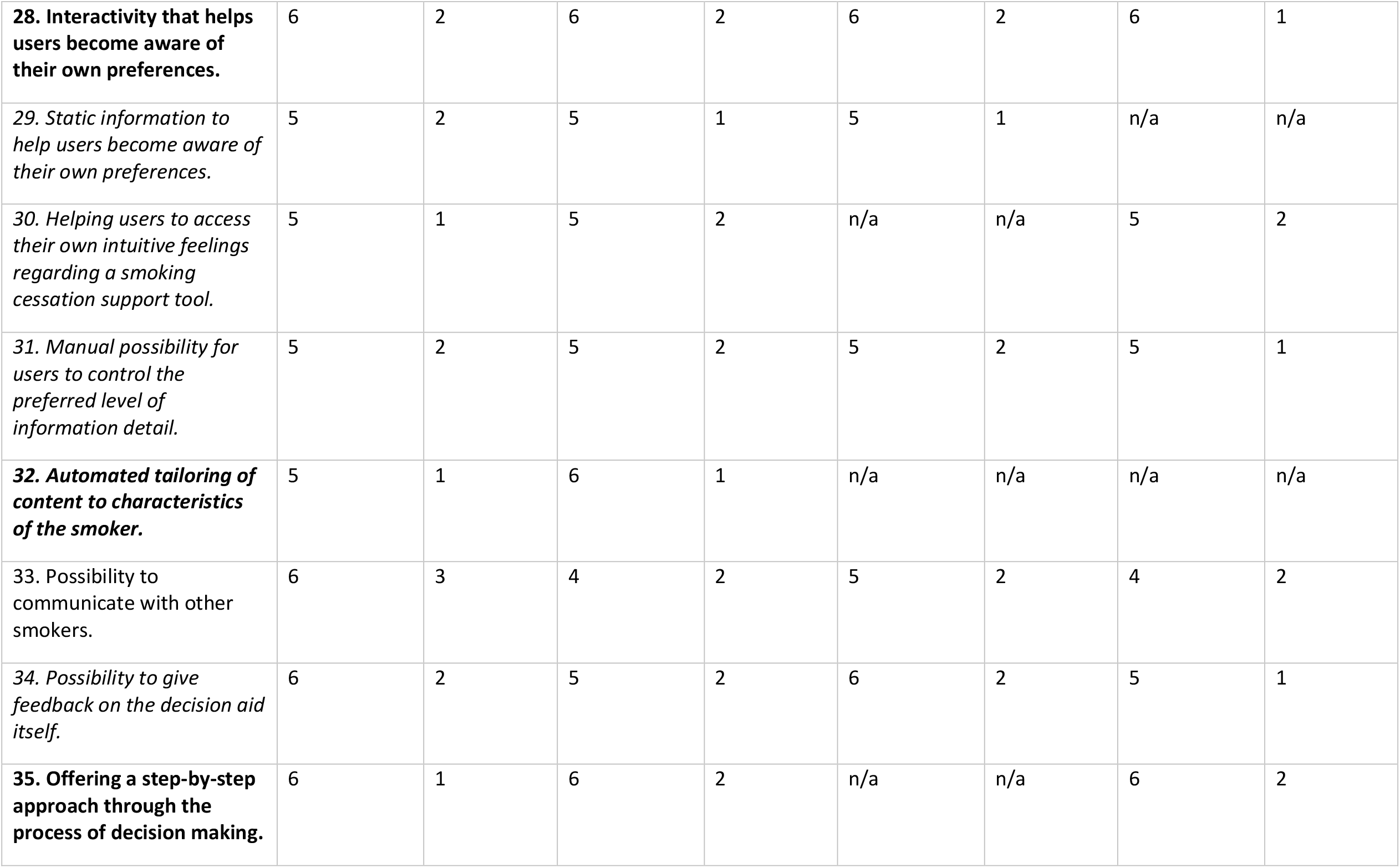
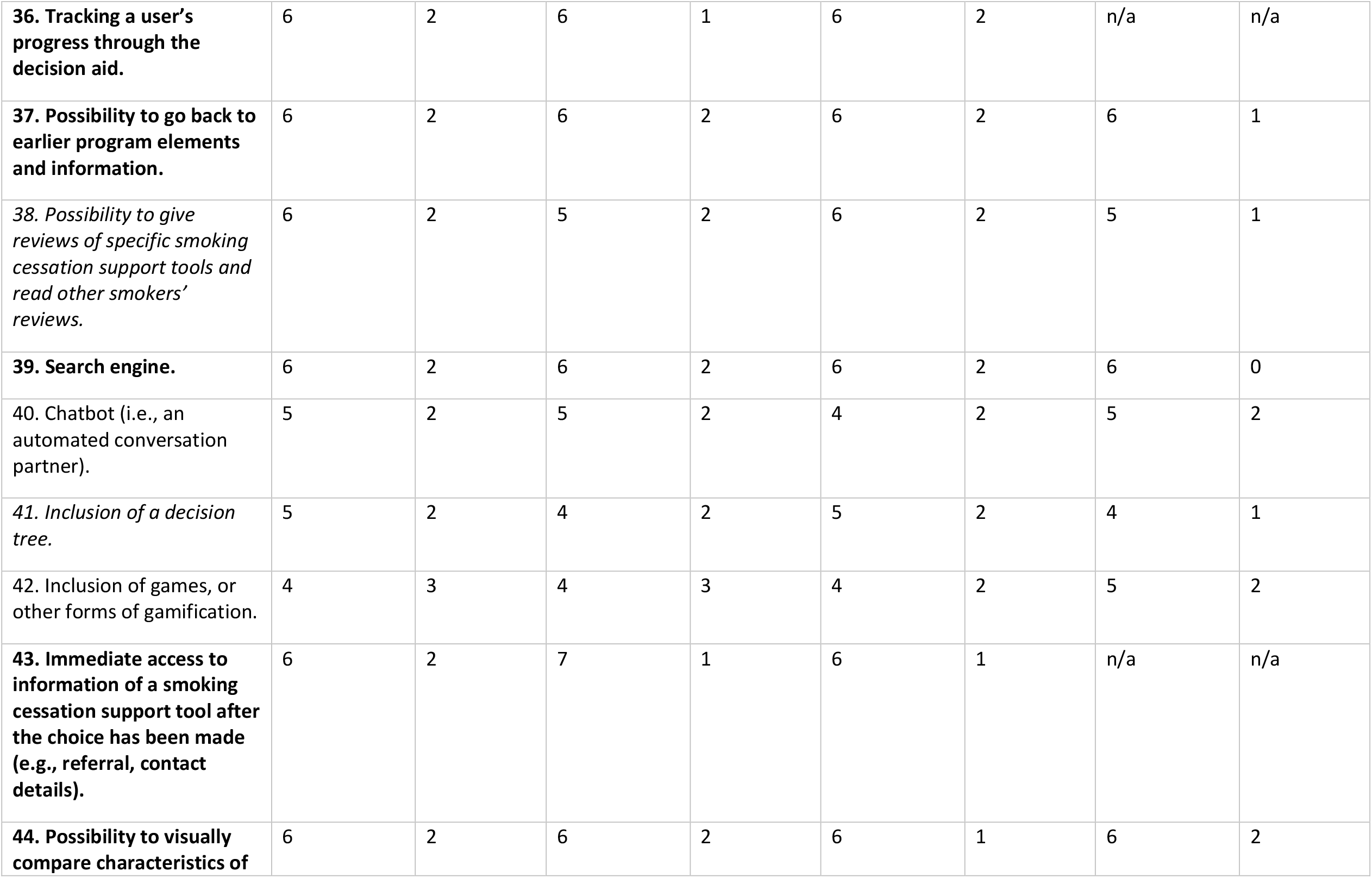
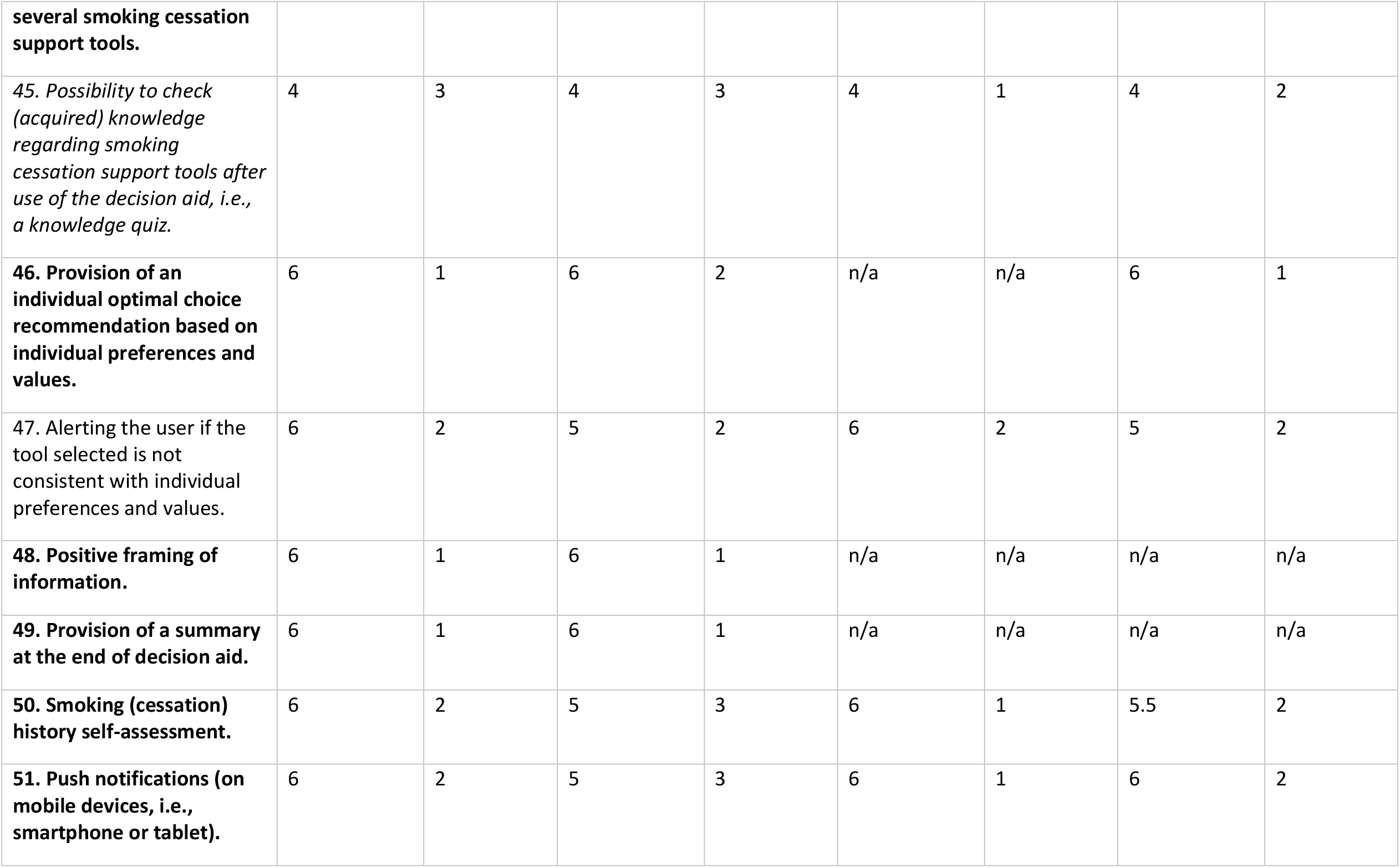
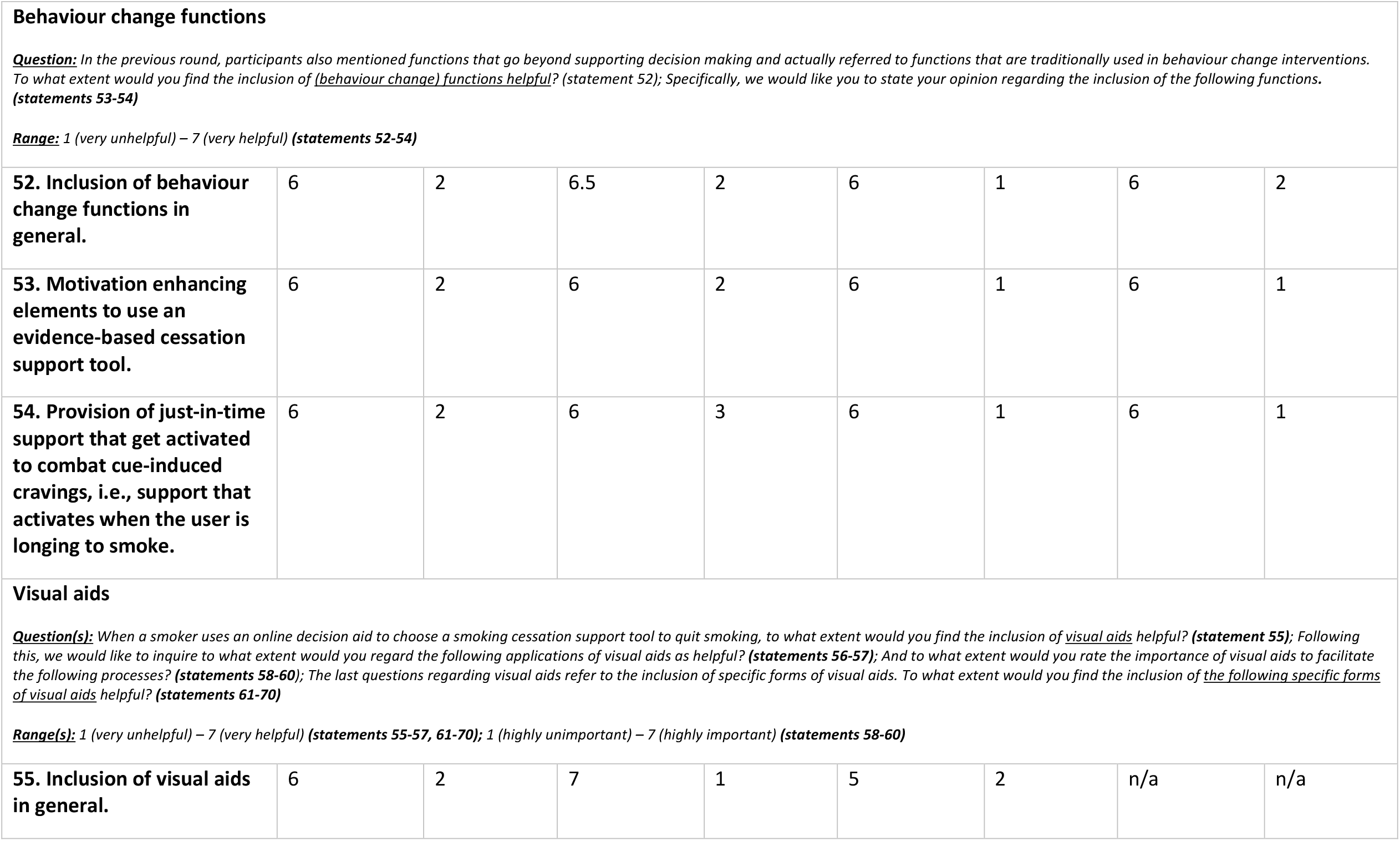
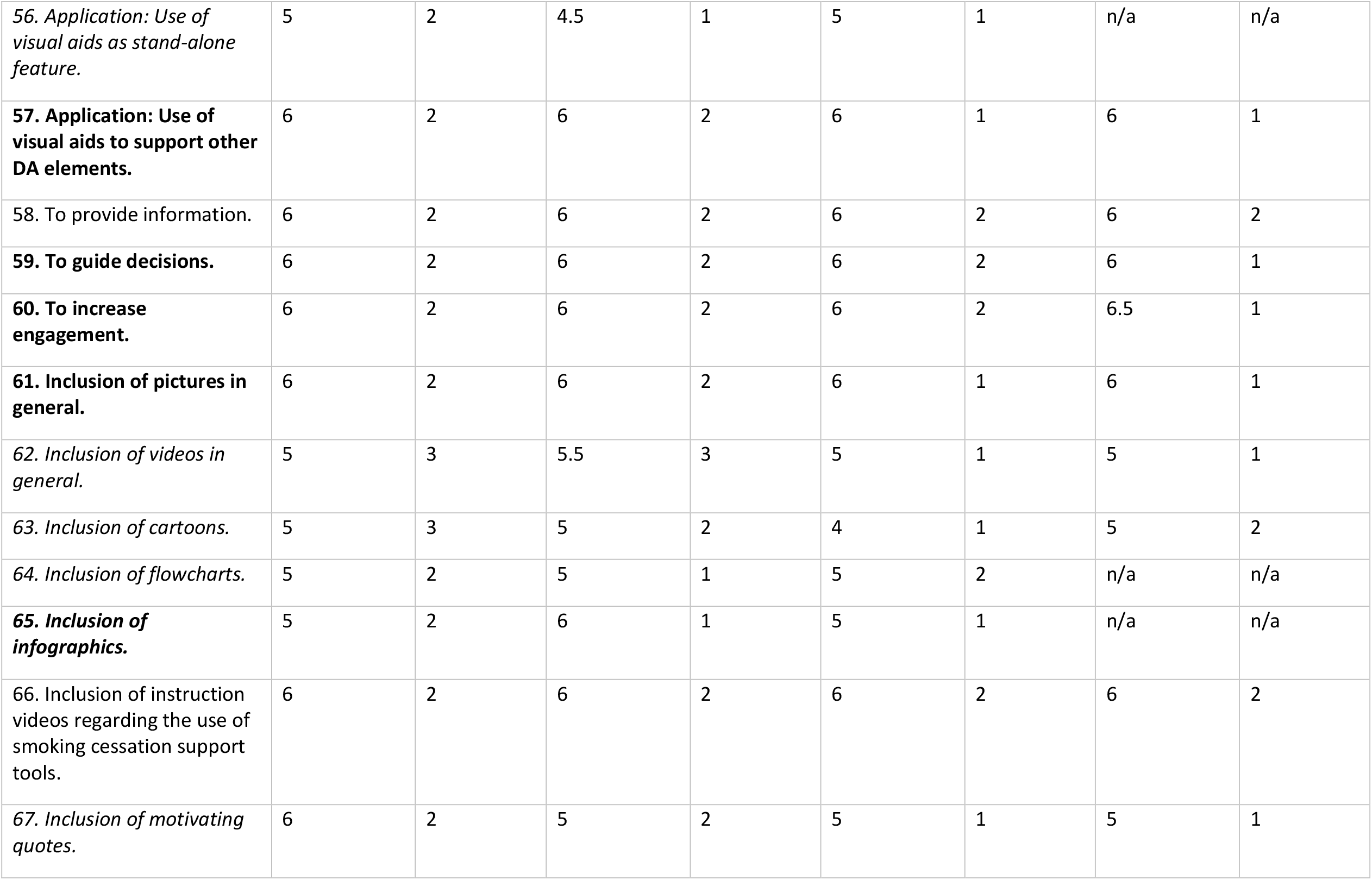
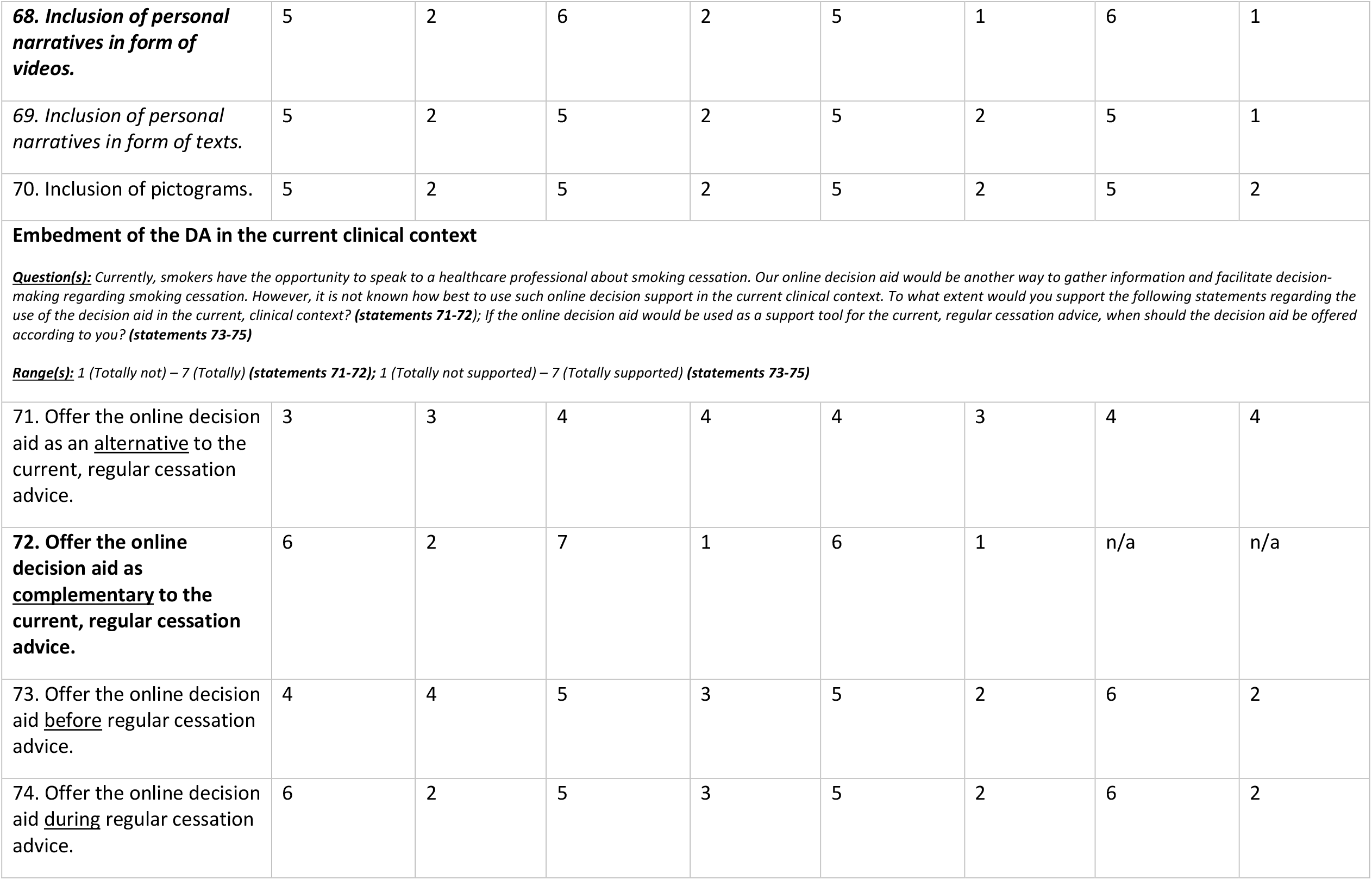
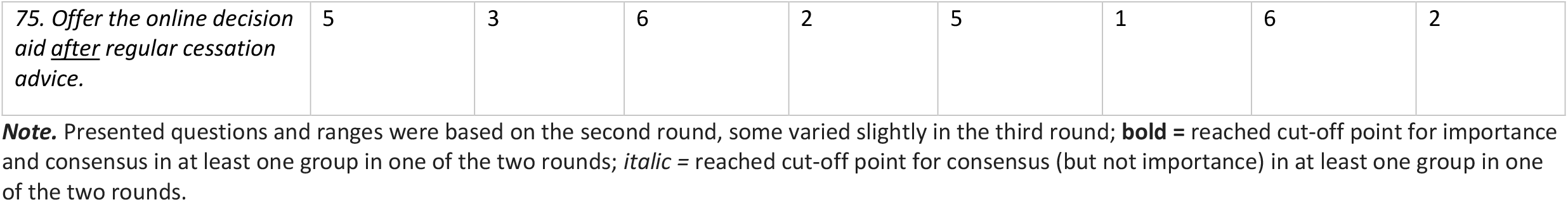
Statements second and third round – importance and consensus.

|  | Second round |  |  |  | Third round |  |  |  |
| --- | --- | --- | --- | --- | --- | --- | --- | --- |
| Statement | Smoking cessation counsellors |  | Scientific experts |  | Smoking cessation counsellors |  | Scientific experts |  |
|  | *Median*<br>*(Importance)* | *IQR*<br>*(Consensus)* | *Median*<br>*(Importance)* | *IQR*<br>*(Consensus)* | *Median*<br>*(Importance)* | *IQR*<br>*(Consensus)* | *Median*<br>*(Importance)* | *IQR*<br>*(Consensus)* |
| **The extent to which the cessation support tool ...**<br>***Question(s):** When a smoker makes a decision regarding using a **smoking cessation support tool** to quit smoking, according to you, how important are the <u>following aspects of cessation support tools</u> in this decision? (statements 1-26)*<br>***Ranges(s):** 1 (highly unimportant) – 7 (highly important) (statements 1-26)* |  |  |  |  |  |  |  |  |
| **1. includes personal guidance during use.** | 6 | 2 | 6 | 2 | 6 | 2 | 6 | 1 |
| **2. is understandable to all smokers (with varying degrees of literacy).** | 7 | 1 | 7 | 1 | n/a | n/a | n/a | n/a |
| **3. is easy to use.** | 6 | 1 | 7 | 0 | n/a | n/a | n/a | n/a |
| **4. requires costs to use.** | 5 | 2 | 4 | 5 | 6 | 1 | 6 | 2 |
| 5. has shown to be effective based on scientific evidence. | 6 | 2 | 6 | 3 | 6 | 2 | 6 | 3 |
| **6. meets the smoker's individual preferences and values for cessation support.** | 6 | 1 | 7 | 2 | n/a | n/a | 7 | 1 |
| 7. is pharmacology-based. | 4 | 2 | 4 | 3 | 4 | 2 | 4 | 2 |
| 8. *is technology-based.* | 4 | 2 | 4 | 2 | 4 | 1 | 4.5 | 2 |
| **9. can be tailored towards the individual user.** | 6 | 2 | 6 | 2 | 6 | 2 | 6 | 1 |
| **10. is appropriate for the user's level of addiction.** | 6 | 1 | 6 | 1 | n/a | n/a | n/a | n/a |
| 11. has certain contraindications. | 6 | 2 | 4 | 2 | 6 | 3 | 4 | 2 |
| 12. has specific usage limitations. | 6 | 3 | 4 | 2 | 5 | 2 | 4 | 2 |
| **13. has (severe) side effects.** | 6 | 2 | 5 | 2 | 6 | 1 | 5 | 2 |
| **14. has a well-described working-mechanism.** | 6 | 2 | 4.5 | 3 | 6 | 1 | 4 | 3 |
| 15. *requires actions from the user.* | 6 | 2 | 5 | 2 | 6 | 2 | 5 | 1 |
| **16. is emotionally appealing to the smoker.** | 6 | 1 | 6 | 2 | n/a | n/a | 6 | 1 |
| 17. takes a certain amount of time. | 5 | 2 | 4.5 | 2 | 5 | 2 | 5 | 2 |
| 18. must be used a number of times. | 5 | 2 | 4 | 3 | 5 | 2 | 4 | 2 |
| **19. is visually appealing to the user.** | 4 | 3 | 6 | 2 | 4 | 2 | 6 | 1 |
| **20. is safe to use.** | 7 | 1 | 6 | 2 | n/a | n/a | 6 | 2 |
| 21. requires travel time to gain access to the tool. | 4 | 3 | 4 | 3 | 5 | 3 | 5 | 2 |
| 22. is tied to a financial incentive. | 4 | 3 | 4 | 3 | 4 | 3 | 4 | 2 |
| **23. is easy to obtain for smokers.** | 6 | 3 | 7 | 1 | 6 | 1 | n/a | n/a |
| **24. involves regular contact with a healthcare professional.** | 6 | 2 | 4 | 2 | 6 | 1 | 4 | 2 |
| 25. is used together with other smokers in a group setting. | 4 | 2 | 4 | 2 | 4 | 2 | 4 | 1 |
| 26. is advised to be used by a healthcare professional. | 5 | 2 | 4.5 | 2 | 5 | 1 | 5 | 2 |
| **Functions of the decision aid**<br><u>**Question(s):**</u> When a smoker uses an online decision aid aimed at facilitating the decision-making process of choosing a smoking cessation support tool to quit smoking, to what extent would you find the inclusion of the following functions helpful? (statements 27-51)<br><u>**Range(s):**</u> 1 (very unhelpful) – 7 (very helpful) (statements 27-51) |  |  |  |  |  |  |  |  |
| 27. Making user aware why a decision has to be made. | 6 | 2 | 5 | 2 | 6 | 2 | 5 | 2 |
| **28. Interactivity that helps users become aware of their own preferences.** | 6 | 2 | 6 | 2 | 6 | 2 | 6 | 1 |
| *29. Static information to help users become aware of their own preferences.* | 5 | 2 | 5 | 1 | 5 | 1 | n/a | n/a |
| *30. Helping users to access their own intuitive feelings regarding a smoking cessation support tool.* | 5 | 1 | 5 | 2 | n/a | n/a | 5 | 2 |
| *31. Manual possibility for users to control the preferred level of information detail.* | 5 | 2 | 5 | 2 | 5 | 2 | 5 | 1 |
| **32. Automated tailoring of content to characteristics of the smoker.** | 5 | 1 | 6 | 1 | n/a | n/a | n/a | n/a |
| 33. Possibility to communicate with other smokers. | 6 | 3 | 4 | 2 | 5 | 2 | 4 | 2 |
| *34. Possibility to give feedback on the decision aid itself.* | 6 | 2 | 5 | 2 | 6 | 2 | 5 | 1 |
| **35. Offering a step-by-step approach through the process of decision making.** | 6 | 1 | 6 | 2 | n/a | n/a | 6 | 2 |
| **36. Tracking a user's progress through the decision aid.** | 6 | 2 | 6 | 1 | 6 | 2 | n/a | n/a |
| **37. Possibility to go back to earlier program elements and information.** | 6 | 2 | 6 | 2 | 6 | 2 | 6 | 1 |
| *38. Possibility to give reviews of specific smoking cessation support tools and read other smokers' reviews.* | 6 | 2 | 5 | 2 | 6 | 2 | 5 | 1 |
| **39. Search engine.** | 6 | 2 | 6 | 2 | 6 | 2 | 6 | 0 |
| 40. Chatbot (i.e., an automated conversation partner). | 5 | 2 | 5 | 2 | 4 | 2 | 5 | 2 |
| *41. Inclusion of a decision tree.* | 5 | 2 | 4 | 2 | 5 | 2 | 4 | 1 |
| 42. Inclusion of games, or other forms of gamification. | 4 | 3 | 4 | 3 | 4 | 2 | 5 | 2 |
| **43. Immediate access to information of a smoking cessation support tool after the choice has been made (e.g., referral, contact details).** | 6 | 2 | 7 | 1 | 6 | 1 | n/a | n/a |
| **44. Possibility to visually compare characteristics of** | 6 | 2 | 6 | 2 | 6 | 1 | 6 | 2 |
| **several smoking cessation support tools.** |  |  |  |  |  |  |  |  |
| *45. Possibility to check (acquired) knowledge regarding smoking cessation support tools after use of the decision aid, i.e., a knowledge quiz.* | 4 | 3 | 4 | 3 | 4 | 1 | 4 | 2 |
| **46. Provision of an individual optimal choice recommendation based on individual preferences and values.** | 6 | 1 | 6 | 2 | n/a | n/a | 6 | 1 |
| 47. Alerting the user if the tool selected is not consistent with individual preferences and values. | 6 | 2 | 5 | 2 | 6 | 2 | 5 | 2 |
| **48. Positive framing of information.** | 6 | 1 | 6 | 1 | n/a | n/a | n/a | n/a |
| **49. Provision of a summary at the end of decision aid.** | 6 | 1 | 6 | 1 | n/a | n/a | n/a | n/a |
| **50. Smoking (cessation) history self-assessment.** | 6 | 2 | 5 | 3 | 6 | 1 | 5.5 | 2 |
| **51. Push notifications (on mobile devices, i.e., smartphone or tablet).** | 6 | 2 | 5 | 3 | 6 | 1 | 6 | 2 |
| **52. Inclusion of behaviour change functions in general.** | 6 | 2 | 6.5 | 2 | 6 | 1 | 6 | 2 |
| **53. Motivation enhancing elements to use an evidence-based cessation support tool.** | 6 | 2 | 6 | 2 | 6 | 1 | 6 | 1 |
| **54. Provision of just-in-time support that get activated to combat cue-induced cravings, i.e., support that activates when the user is longing to smoke.** | 6 | 2 | 6 | 3 | 6 | 1 | 6 | 1 |
| **55. Inclusion of visual aids in general.** | 6 | 2 | 7 | 1 | 5 | 2 | n/a | n/a |
| *56. Application: Use of visual aids as stand-alone feature.* | 5 | 2 | 4.5 | 1 | 5 | 1 | n/a | n/a |
| **57. Application: Use of visual aids to support other DA elements.** | 6 | 2 | 6 | 2 | 6 | 1 | 6 | 1 |
| 58. To provide information. | 6 | 2 | 6 | 2 | 6 | 2 | 6 | 2 |
| **59. To guide decisions.** | 6 | 2 | 6 | 2 | 6 | 2 | 6 | 1 |
| **60. To increase engagement.** | 6 | 2 | 6 | 2 | 6 | 2 | 6.5 | 1 |
| **61. Inclusion of pictures in general.** | 6 | 2 | 6 | 2 | 6 | 1 | 6 | 1 |
| *62. Inclusion of videos in general.* | 5 | 3 | 5.5 | 3 | 5 | 1 | 5 | 1 |
| *63. Inclusion of cartoons.* | 5 | 3 | 5 | 2 | 4 | 1 | 5 | 2 |
| *64. Inclusion of flowcharts.* | 5 | 2 | 5 | 1 | 5 | 2 | n/a | n/a |
| **65. Inclusion of infographics.** | 5 | 2 | 6 | 1 | 5 | 1 | n/a | n/a |
| 66. Inclusion of instruction videos regarding the use of smoking cessation support tools. | 6 | 2 | 6 | 2 | 6 | 2 | 6 | 2 |
| *67. Inclusion of motivating quotes.* | 6 | 2 | 5 | 2 | 5 | 1 | 5 | 1 |
| 68. Inclusion of personal narratives in form of videos. | 5 | 2 | 6 | 2 | 5 | 1 | 6 | 1 |
| 69. Inclusion of personal narratives in form of texts. | 5 | 2 | 5 | 2 | 5 | 2 | 5 | 1 |
| 70. Inclusion of pictograms. | 5 | 2 | 5 | 2 | 5 | 2 | 5 | 2 |
| **Embedment of the DA in the current clinical context**<br>***Question(s):** Currently, smokers have the opportunity to speak to a healthcare professional about smoking cessation. Our online decision aid would be another way to gather information and facilitate decision-making regarding smoking cessation. However, it is not known how best to use such online decision support in the current clinical context. To what extent would you support the following statements regarding the use of the decision aid in the current, clinical context? (statements 71-72); If the online decision aid would be used as a support tool for the current, regular cessation advice, when should the decision aid be offered according to you? (statements 73-75)*<br>***Range(s):** 1 (Totally not) – 7 (Totally) (statements 71-72); 1 (Totally not supported) – 7 (Totally supported) (statements 73-75)* |  |  |  |  |  |  |  |  |
| 71. Offer the online decision aid as an <u>alternative</u> to the current, regular cessation advice. | 3 | 3 | 4 | 4 | 4 | 3 | 4 | 4 |
| 72. Offer the online decision aid as <u>complementary</u> to the current, regular cessation advice. | 6 | 2 | 7 | 1 | 6 | 1 | n/a | n/a |
| 73. Offer the online decision aid <u>before</u> regular cessation advice. | 4 | 4 | 5 | 3 | 5 | 2 | 6 | 2 |
| 74. Offer the online decision aid <u>during</u> regular cessation advice. | 6 | 2 | 5 | 3 | 5 | 2 | 6 | 2 |
| 75. Offer the online decision aid *after* regular cessation advice. | 5 | 3 | 6 | 2 | 5 | 1 | 6 | 2 |

**Note.** Presented questions and ranges were based on the second round, some varied slightly in the third round; **bold** = reached cut-off point for importance and consensus in at least one group in one of the two rounds; *italic* = reached cut-off point for consensus (but not importance) in at least one group in one of the two rounds.

The questionnaire of the third round included only those statements that smoking cessation counsellors and scientific experts failed to reach consensus on in the second round (63 and 60 statements, respectively). Participants received feedback on how much experts agreed with the statements (importance) and each other (consensus).

Questionnaires for the smoking cessation counsellors were formulated in Dutch, while the questionnaires for the scientific experts were formulated in English.

#### Data analysis

The data of the first rounds were analysed using NVivo 12 (QSR International, 2018). Again, TG and RH created a code tree in an iterative process and used it to code all answers separately and reached a substantial intercoder reliability of 0.67 (Cohen’s Kappa) and 99.24% (percentage of agreement).

The data of the second and third rounds were analysed with SPSS (IBM Corp, 2017). Respondents’ answers were included if respondents answered at least 80% of the questions. Frequencies were used to examine the demographics; medians were calculated to examine which importance participants (as a group) attributed to individual statements (≥6 was regarded as cut-off point) and interquartile ranges (IQR) were used to examine the consensus reached for every statement (≤1 was regarded as cut-off point). Cut-off points were based on similar studies (e.g., Schneider et al., 2012). The two expert groups were analysed separately.

## Results

### In-depth semi-structured interviews

Twenty individual interviews were conducted. Nine (45%) of the interviewees identified as women, while the other 11 (55%) identified as men. The majority (n=12, 60%) held a medium level of education and on average interviewees undertook three previous smoking cessation attempts that lasted for 178 days on average *(Table 2)*.

**Table 2.** Descriptive characteristics of participants (N=20)

|  |  |  |
| --- | --- | --- |
| **Mean age in years (SD)** | 50.3 | (13.5) |
| **Gender** |  |  |
| **Identified as women (%)** | 9 | (45%) |
| **Identified as men (%)** | 11 | (55%) |
| **Identified as another gender/non-binary (%)** | 0 | (0%) |
| **Education** |  |  |
| **Low (%)**<br>*No, primary and vocational education* | 1 | (5%) |
| **Medium (%)**<br>*Secondary vocational education and a high school degree* | 12 | (60%) |
| **High (%)**<br>*Higher vocational education, college and university degrees* | 7 | (35%) |
| **Smoking behaviour** |  |  |
| **Mean number of quit attempts (SD)** | 3.0 | (2.1) |
| **Mean duration of quit attempt in days (SD)** | 177.8 | 445 |
| **Mean intention to use the decision aid (SD)\*** | 4.2 | 2.0 |

**Note.** \*Intention: 1 = low intention; 7 = high intention

### Knowledge

Overall, the level of cessation assistance tool knowledge varied considerably between individuals. Interviewees tended to be aware of some facts (e.g., tool’s side-effects) without being aware of other facts (e.g., insurance reimbursement). The terms (non-)evidence-based also were not always clear, and some interviewees were not aware of the distinction between evidence-based and non-evidence-based tools. Interviewees also sometimes equated believing and knowing, for example by expressing that they did not *believe* in the distinction between evidence-based and non-evidence-based tools. Interestingly, some interviewees displayed a discrepancy between perceived and objective knowledge, i.e., they regarded themselves as being informed while also lacking information.

Multiple information sources were described to acquire knowledge: (1) Other people’s personal experiences, such as colleagues or neighbours; (2) one’s own previous experience(s); and (3) the media. Sometimes interviewees described shortcoming of these strategies without necessarily realising them, e.g., someone considered an evidence-based tool to be ineffective *in general* because it was ineffective *for them personally*; and a few people described that one source was more important than the other, e.g., personal experiences as opposed to other people’s experiences.

### Values related to cessation assistance

Values and beliefs were notably very heterogenous and often linked to personal circumstances. Interviewees also described aspects that directly influenced decision making for them that were not inherent to the tools, e.g., one’s personal experiences with a tool. Despite the variety, one pattern emerged: Effectiveness was *referred* to the most, followed by costs. The same pattern also emerged when the interviewees were asked about the characteristics that were the most important to them. Interviewees often made references to long-term success and some used the term in a broader way (e.g., a tool has to fit personally in order to be effective). Also, valuing effectiveness as important did not seem to be related to educational level, although this idea was conveyed by one of the highly educated interviewees. Other particularly important values were: (1) That the tool is covered by health Insurance; (2) that the tool is offered by a(n) (professional) expert (e.g., general practitioners); (3) side effects; (4) safety; (5) personal contact with another person; (6) personal fit; and (7) practical considerations, such as length of treatment, number of doses/sessions and the location. The only characteristic that was almost universally accepted as being unimportant was whether tools were advertised. Other characteristics were alternately framed as advantages or disadvantages, e.g., some interviewees appreciated personal contact, while others did not. Interviewees also clearly made trade-offs during the interviews, e.g., by describing that certain characteristics were important but less so than others. For example, short-term side effects that were considered as unimportant and long-term side effects that were considered as important. Temporal dynamics of values were also acknowledged by describing that the evaluations of certain characteristics depend on the smoking cessation phase one is in, e.g., one interviewee indicated that personal contact would become important after the period in which they were able to *not* smoke on their own.

### DA

Interviewees often referred to the materials we sent before the interview; those were regarded as pleasant or useful and interviewees appreciated that the differences between the different cessation tools became apparent by naming the tools’ advantages and disadvantages – this could be used to compare tools to make a decision. Also, some interviewees seemed to expect that they would receive advice from the DA. The majority of the interviewees were at least somewhat interested in using the proposed DA. The interviewees that were either not interested or less interested in the DA had different reasons, e.g., mistrust in online information, that the information could also be acquired somewhere else (e.g., through a face-to-face meeting with a healthcare professional) or not being interested in cessation (assistance) at the moment. Other interviewees did not fully grasp the concept of a DA at the beginning and needed more explanation. No clear patterns emerged based on gender identity, age and level of education – however, fewer younger interviewees (<40) talked about mistrust in online information or that information could be acquired elsewhere.

### Personal needs regarding functional aspects of the DA

Interviewees often indicated that they liked the table with all options that we presented as preparation for the interviews. A number of interviewees also indicated that they would like to hear about others’ choices and experiences. The content of the offered information was discussed as well, with information that often related to earlier described values (see *Values related to cessation assistance*). Interestingly, no-one indicated that the inclusion of certain functions would make the DA uninteresting for them, rather they indicated that they would skip the function or that they would use the function passively. Functions were often linked to personal circumstances and/or preferences. Overall, interviewees often liked the idea that they could have control over the amount of information in the DA.

### Personal needs regarding device compatibility & usage duration of the DA

Multiple possible devices (e.g., personal computers) to use the DA were named, as were various time spans. The majority wanted to use the DA on their computer, and only a few mentioned mobile phones – only one interviewee wanted to use the DA as a mobile phone app. Interestingly, no-one from the oldest age group (>69) and only one person from the age group 30-39 indicated that they would like to use the DA on their mobile phone and people with a medium level of education seemed to prefer mobile phones more.

Preferred time spans ranged from 5 minutes to an unlimited time span, however, some were unable to name a time span at all. Older interviewees (>50) did not indicate a clear time frame, with one interviewee (70-year-old) stressing the amount of time at their disposal. A few interviewees who preferred short time spans indicated that they would be willing to spend more time if they were in doubt or if they wanted to receive extra information. Some interviewees indicated that they would want to use the DA multiple times.

### Personal needs regarding the layout of the DA

Interviewees overall indicated that they would like the DA to be neutral, functional, simple, positive and motivating, however quite a few interviewees found the question difficult to answer. A number of interviewees expressed that they rather not see any negative or off-putting imagery, such as used on cigarette packs. Regarding visual aids, some interviews indicated that they could be supportive but should only be used if they were of added value (e.g., pictures of different tools to recognise them), while others indicated that they would not be needed for them at all. People who disfavoured visual aids tended to identify as women between 40-59 (most 40-49) years old with a medium level of education. Overall, an intuitive and easy to navigate layout seemed to be preferred.

## Three-round Delphi study

### First round

Nine scientific experts and eight smoking cessation counsellors participated in the first round. The majority of the scientific experts were either associate (n=1, 11.1%) or full professors (n=4, 44.4%), while the biggest group of the smoking cessation counsellors were practice nurses (n=6, 75%). Answers fell into nine categories *(Table 3)*.

**Table 3.** Answer categories and examples of the first round.

|  | Answer categories | Answer examples |
| --- | --- | --- |
| 1. | Advertising the decision aid | If a patient comes to a GP for a consultation [...] point out this possibility [the decision aid] to the patient. |
| 2. | Characteristics of smoking cessation assistance tools | Effectiveness |
| 3. | Characteristics of users | Previous cessation attempts and experience |
| 4. | Content of the decision aid regarding evidence-based cessation tools | If it includes non evidence-based tools this has to be clearly stated |
| 5. | Decision aid attributes | Accessibility |
| 6. | Decision aid functions | Possibility to obtain in-depth info when needed |
| 7. | Layout considerations | Match with target audience |
| 8. | Use of the decision aid in the current clinical context | Healthcare providers could show it [the decision aid] to patients and they can use the decision aid together. |
| 9. | Other | Actors need to be appealing to the end-users. Tip: Involve the end-user in the development of the tool and especially in the development of the visual aids to |
|  |  | ensure these appeal to them and match their needs and wants. |

**Note.** GP = general practitioner

### Second and third round

#### Sample

Sixty-one smoking cessation counsellors and 44 scientific experts participated in the second round; 62.3% (n=38) of the smoking cessation counsellors and 54.5% of the scientific experts (n=24) returned to fill in the third round. In both rounds, the majority of the smoking cessation counsellors were practice nurses in both the second (n=43, 70.5%) and third round (n=29, 76.3%), while the majority of the scientific experts were either associate (n=9, 20.5% in the second round; and n=4, 16.7% in the third round) or full professors (n=16, 36.4% in the second round; and n=8, 33.3% in the third round). More details regarding the samples of the second and third rounds can be found in *Appendix 1*.

### Consensus and importance

The smoking cessation counsellors reached consensus (IQR ≤1) on 12 statements (16% of all statements) in the second round, and on 26 statements (35% of all statements, 41% of all statements in the third round) in the third round. The scientific experts reached consensus (IQR ≤1) on 15 statements (20% of all statements) in the second round, and on 25 statements (33% of all statements, 42% of all statements in the third round) in the third round. See *Table 4* for all statements and the respective scores for importance and consensus.

## Discussion

The aim of this study was to explore the needs and viewpoints regarding supporting informed decision making within the context of smoking cessation from the perspective of three stakeholders: (1) Potential end users (i.e., individuals motivated to quit smoking), (2) smoking cessation counsellors and (3) scientific experts.

### Potential end users (i.e., individuals motivated to quit smoking)

Most importantly the interview study showed that potential end users are not a completely homogenous group – especially not in terms of knowledge, but also to a certain degree in terms of values or when queried about the desired characteristics of a potential DA. We will reflect on all three aspects in the following.

Especially the first two aspects (i.e., heterogeneity in knowledge and values) highlight that DAs might be valuable interventions to support individuals during smoking cessation attempts, as their two core elements–information provision (Abhyankar et al., 2013) and so-called value clarification methods (Fagerlin et al., 2013)–are designed to support users in acquiring more knowledge and using this knowledge to identify what is important to them personally, and have shown to be effective in doing just that (Stacey et al., 2017). Our results indicate that individuals motivated to quit smoking especially regard the effectiveness of smoking cessation assistance as important and, therefore, should be provided with information about this aspect of said assistance. Interestingly, however, these end users seemingly understand the concept of effectiveness differently from experts. This becomes clear when one looks at the way end users acquire and interpret information around effectiveness. For example, end users consider smoking cessation assistance to be effective if someone in their environment has stopped while using assistance and consider a tool to be ineffective if they themselves have used it once and ultimately have not quit smoking. This indicates that laypersons understand information about effectiveness on a personal or individual level, while scientists interpret the effectiveness of a tool by comparing different groups (e.g., one group receives an evidence-based smoking cessation assistance tool, while the other group receives ‘care-as-usual’) and examining whether additional effects arise on top of what you would expect anyway. Therefore, evidence-based tools are not 100 percent effective, but simply more effective than the comparator (e.g., ‘care-as-usual’). Laypeople motivated to quit smoking do not seem to be aware of this crucial information possibly leading them to rather not use an evidence-based tool in the future – thereby, they unknowingly decrease their own cessation chances. It therefore seems important to provide end users with accurate information regarding effectiveness in an accessible manner to prevent them from not using a specific tool or evidence-based tools in general after a failed quit attempt with smoking cessation assistance. One way to achieve this could be to use icon arrays. These are graphical representations consisting of a number of icons that symbolize individuals within a group who are affected by a certain event (Galesic et al., 2009). Such icon arrays are mostly used to convey risk information and have been shown to increase accuracy of understanding (Galesic et al., 2009). Based on the results from the needs assessment described in this paper and on previous findings from risk communication studies, we therefore decided to make icon arrays for our DA (using the website Iconarray.com (Risk Science Center and Center for Bioethics and Social Sciences in Medicine, University of Michigan, n.d.)) to convey information regarding the effectiveness of different smoking cessation assistance tools. More information on the application of this method can be found in our paper describing the DA (Gültzow, Smit, Hudales, Knapen, et al., 2020).

The third aspect that made the end user group rather heterogenous (i.e., varying needs regarding DA characteristics) shows that smoking cessation DAs should be more flexible in terms of functionalities to appeal to a wide audience and should be adaptable to–and potentially by–the end users themselves. This most likely means that digital DAs should be offered to individuals wishing to quit smoking as digital tools allow for more adaptability (Hoffman et al., 2013). Health psychologist and health promoters often acknowledge two different forms of adaption to users characteristics: (1) System-driven tailoring, or the automatized adaption of intervention materials to individual users’ characteristics (de Vries & Brug, 1999) and (2) customization, or enabling users to adapt the intervention materials themselves (i.e., via a non-automated process) (Bol et al., 2019). While system-driven tailoring has shown promising results in smoking cessation interventions (e.g., Wangberg et al., 2011), theoretical (Ryan & Deci, 2008) and empirical insights (Syrowatka et al., 2016) suggest that customization is better suited for DAs compared to system-driven tailoring. In fact, allowing users the possibility to customize DAs to a certain degree may lead to greater chances for autonomous behaviour change (Bol et al., 2019). However, developers have to ensure that ‘core functions’ and the most important information that the DA aims to convey should be retained. We ourselves have used various options to deal with this in the smoking cessation DA developed as part of the associated broader project (Gültzow, Smit, Hudales, Knapen, et al., 2020), e.g., we have chosen to enable users (1) to receive in-depth information regarding options that they are interested in *while* ensuring that all users receive the necessary information to make a decision, and (2) to skip certain functionalities that are not regarded as necessary but as attractive by *some* users (e.g., a knowledge quiz).

### Smoking cessation counsellors and scientific experts

The two Delphi studies resulted in a breadth of information that can be used to support DA development for smoking cessation or other preventive health-related behaviours. Interestingly, the two groups only disagreed on a few statements during the last two rounds, i.e., the adoption of (system-driven) tailoring strategies and the inclusion of infographics and personal narratives in the form of videos were deemed important by the scientific experts, but not by the smoking cessation counsellors. This could reflect the composition of the group of scientists whose expertise has been mainly behavioural intervention (development), i.e., experts who often apply behaviour change techniques, such as system-driven tailoring. Here it should be acknowledged that the statements that the experts rated were mostly global in nature, i.e., they mostly referred to smoking cessation DAs overall and not to specific section of DAs. It is also possible to integrate experts’ viewpoints and use tailoring strategies for different purposes, while not using them throughout the entire DA. To illustrate, tailoring in general has been shown to negatively impact on quality of decision making when used in digital DAs (Syrowatka et al., 2016). On the other hand, very specific tailoring in the form of tailored advice based on identified values might help DA-users to make value-consistent choices (Witteman et al., 2020). In our own DA, we therefore aimed for a synergistic approach: Throughout the majority of the DA users were able to customize the information they were receiving (i.e., we used customization as opposed to system-driven tailoring as described in *Potential end users (i*.*e*., *individuals motivated to quit smoking*)), but we also provided them with system-tailored advice at the end (Gültzow, Smit, Hudales, Knapen, et al., 2020). This shows that it is possible to integrate dissenting expert opinions, in future studies it should be assessed if this approach is effective as well.

It should also be noted that both expert groups agreed on a number of statements, e.g., that cessation assistance should be appropriate for users’ level of addiction. This specific statement can also help to illustrate how the information from these two Delphi studies can be used to inform DA development: Developers, often scientific experts, of a smoking cessation DA could include a smoking assessment to provide end users with information about which tools are appropriate for their level of addiction – a function which has also been deemed helpful by the smoking cessation counsellors. Again, this is something we have implemented in our own DA (Gültzow, Smit, Hudales, Knapen, et al., 2020): End users were asked to indicate if they smoke more than 10 cigarettes a day or if they had multiple unsuccessful smoking cessation attempts in the past. Subsequently, end users that showed these characteristics were advised to use a combination of both behavioural as well as pharmacological cessation assistance – as indicated by Dutch smoking cessation guidelines (Chavannes et al., 2017). Another example would be that both expert group groups agreed that smoking cessation assistance should meet individuals’ preferences and values for cessation support. Which is why we are currently testing if a smoking cessation DA is more effective if users are supported in identifying smoking cessation assistance tools that reflect their values and preferences (Gültzow, Smit, Hudales, Knapen, et al., 2020).

### Integration of the results in different stakeholders

Our results show that it is of great importance to include multiple stakeholders during the needs assessment, as different stakeholders’ opinions do not always align; e.g., important values that were identified among potential end users differed from the one’s identified among (professional) experts. That being said, one should always consider which stakeholders should be included. The original IPDAS development process advocates to include both potential end users as well as clinicians as the main focus of the IPDAS collaboration is on DAs that are used within clinical encounters (Coulter et al., 2013). The studies presented in this article show that the inclusion of other stakeholder groups is potentially beneficial for other types of DAs, such as DAs with a health promotion focus. Future DA developers should therefore consider which stakeholders are of importance for their specific DA and not ‘simply’ follow the IPDAS advice. In some cases, one might even consider not to include clinicians, e.g., regarding the decision whether to eat more fruit. Including too many stakeholder groups might overcomplicate things and, for instance, create too many issues about which no consensus can be reached, as illustrated by our finding that different stakeholders’ opinions do not always align.

This is illustrated most clearly when one looks at the aspect of cessation assistance’s effectiveness: Potential end users clearly valued information on effectiveness and largely agreed with each other, while experts failed to establish consensus regarding the value of this type of information. This discrepancy may be due to experts’ and laypersons’ different conceptualizations of the ‘effectiveness’ concept as explained earlier – see *Potential end users (i*.*e*., *individuals motivated to quit smoking*). Other researchers have shown that individuals motivated to quit smoking often do not choose to use smoking cessation assistance tools that have shown to be effective (Cokkinides et al., 2005). Our interviews may give proof to the claim that this might not be due to a disinterest in effectiveness, but that they simply lack the knowledge on how experts conceptualize the term effectiveness. Providing them with this information in an accessible manner (as explained earlier) might thus enable them to choose tools with a greater effectiveness. This specific example shows that integrating the views of different stakeholders can be difficult at first but can also generate valuable insights and that there is no single approach to integrate all discrepancies. In this specific case, it could be sensible to meet the clear informational needs of the end users, who clearly indicated that effectiveness is very important to them. In other cases (e.g., which other characteristics to include in a DA), however, it may be better to take a more balanced approach and combine the insights of the different stakeholder groups. For example, aside from the ‘effectiveness-factor’, we have mostly chosen to only include information when more than one stakeholder group indicated the importance of a factor or used the information provided by the end users to deepen the information presented in the smoking cessation DA that was developed by our team (Gültzow, Smit, Hudales, Knapen, et al., 2020).

### Strengths and limitations

Including three different groups of participants in this needs assessment provided us with a breath of information that can be used to inform the development of smoking cessation DAs. Also, were we generally able to include a diverse group of participants. However, we were unable to include end users identifying as non-binary. Given that gender specifically was not the focus of this study we do not regard this as a *major* issue, yet future studies could make extra efforts to include non-binary individuals, especially because gender and sexuality minority groups are assumed to be provided with less possibilities to engage in high-quality decision making (DeMeester et al., 2016). Likewise, respondents in the group of the smoking cessation counsellors mainly were practice nurses. Yet, as practice nurses increasingly provide smoking cessation counselling in the Netherlands (Freund et al., 2015; Genootschap & Vereniging, 2011), we believe our sample still accurately reflects the viewpoints of counsellors in our Dutch context. We also acknowledge that it could be viewed as a limitation that only Dutch smoking cessation counsellors were involved. However, we assume that our results could be broadly generalizable for other Western countries as Western countries often share similarities in healthcare systems (e.g., the Dutch healthcare systems shows similarities to other Central and Northern European countries, such as the United Kingdom (Ferreira et al., 2018)). Finally, due to the two different study designs and because we treated the two Delphi studies as separate studies (especially after the first round), results are not directly comparable. However, the different study designs and not being able to directly compare the Delphi studies also reflect the choices that we made during the process of the studies to choose a study design that was most fitting for the relevant stakeholder groups and because we preferred to provide the experts only with feedback from their specific peers. That being said, it is technically possible to include end users in Delphi studies as well (e.g., Schneider et al., 2012) and future studies *could* attempt to do this in order to directly compare needs and viewpoints of the different groups which could facilitate comparative studies. At this point, however, it is unclear if this would be of any added value.

## Conclusion

The results from the three studies reported in this article complement the general DA literature and can be used as input for smoking cessation DA development. In fact, we have used the results to develop a digital smoking cessation DA (Gültzow, Smit, Hudales, Knapen, et al., 2020). Given the variation in the needs and wishes revealed among different stakeholders, the combination of these studies highlights that a ‘one size fits all’ approach is not desirable. In the development of future DAs, heterogeneity should certainly be taken into account, for example by enabling users to customize a DA based on their personal preferences while safeguarding essential elements.

## Data Availability

Due to the qualitative nature of the data reported in this article, we have decided not to make the data publicly available.

## Abbreviations

DA: Decision Aid
IPDAS: International Patient Decision Aid Standards
OSF: Open Science Framework

## Acknowledgements

We would like to thank all participants, and Dr. Francine Schneider who advised on the design of the questionnaire for the third round of the Delphi study in order to optimally present the results of the second round to the experts. Additionally, we would like to thank Yil Severijns who provided feedback on parts of the article.

---

**Appendix 1.**
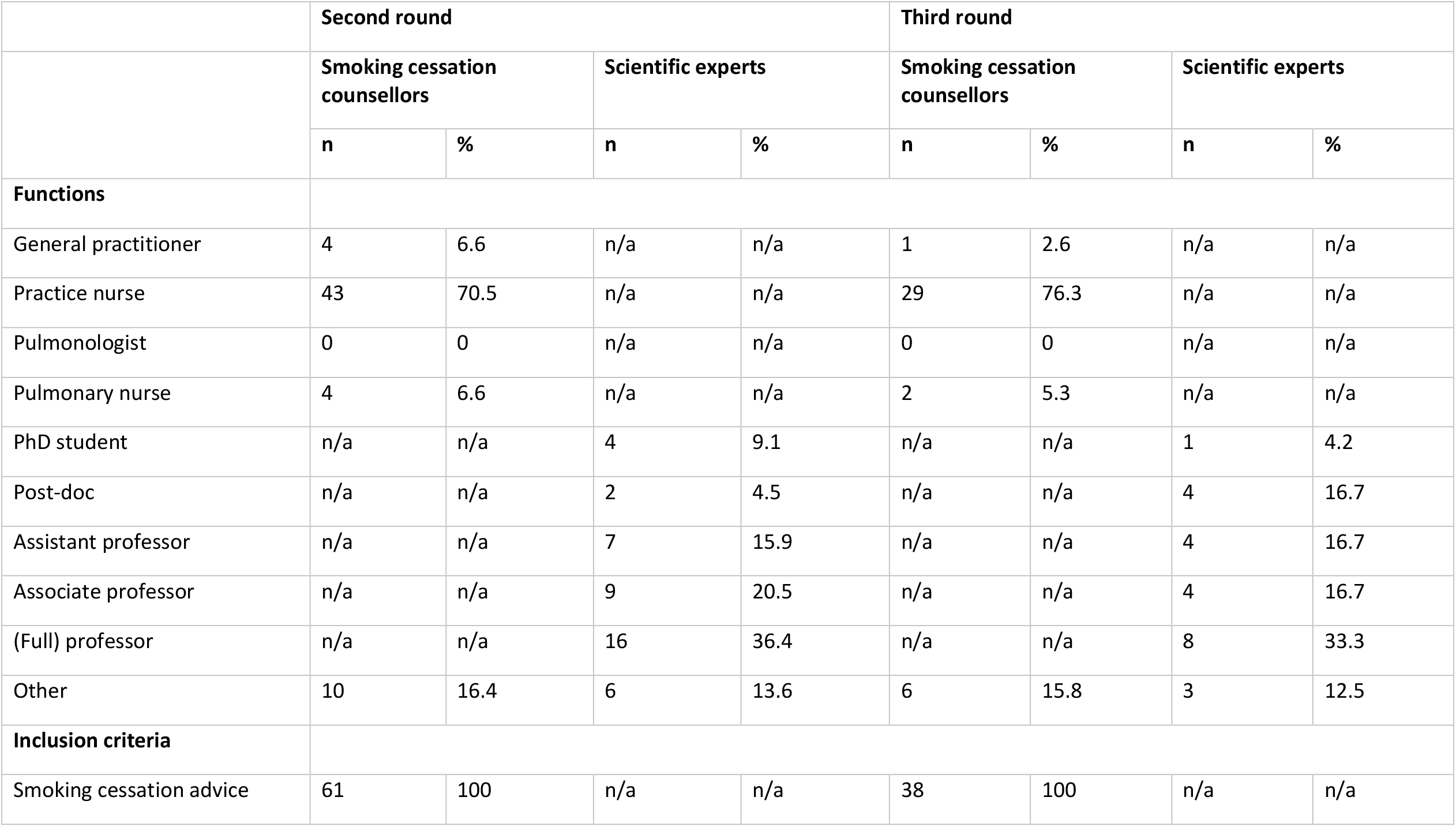
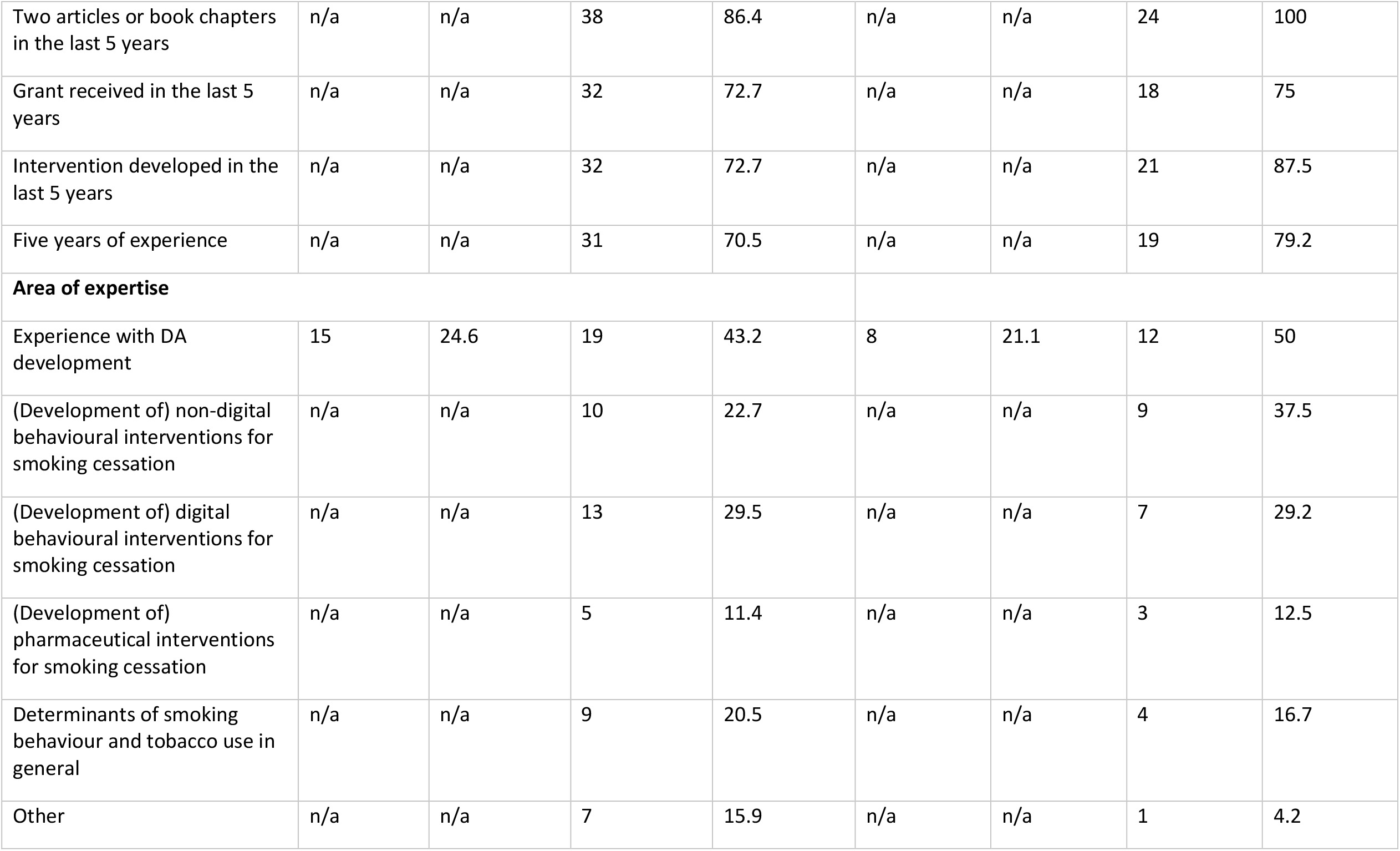
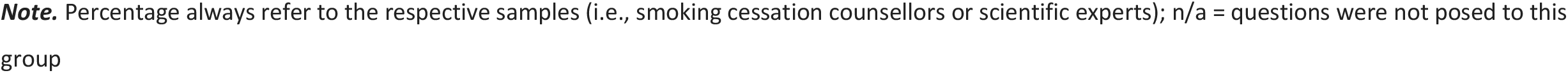
Sample characteristics second and third round.

|  | Second round |  |  |  | Third round |  |  |  |
| --- | --- | --- | --- | --- | --- | --- | --- | --- |
|  | Smoking cessation counsellors |  | Scientific experts |  | Smoking cessation counsellors |  | Scientific experts |  |
|  | n | % | n | % | n | % | n | % |
| **Functions** |  |  |  |  |  |  |  |  |
| General practitioner | 4 | 6.6 | n/a | n/a | 1 | 2.6 | n/a | n/a |
| Practice nurse | 43 | 70.5 | n/a | n/a | 29 | 76.3 | n/a | n/a |
| Pulmonologist | 0 | 0 | n/a | n/a | 0 | 0 | n/a | n/a |
| Pulmonary nurse | 4 | 6.6 | n/a | n/a | 2 | 5.3 | n/a | n/a |
| PhD student | n/a | n/a | 4 | 9.1 | n/a | n/a | 1 | 4.2 |
| Post-doc | n/a | n/a | 2 | 4.5 | n/a | n/a | 4 | 16.7 |
| Assistant professor | n/a | n/a | 7 | 15.9 | n/a | n/a | 4 | 16.7 |
| Associate professor | n/a | n/a | 9 | 20.5 | n/a | n/a | 4 | 16.7 |
| (Full) professor | n/a | n/a | 16 | 36.4 | n/a | n/a | 8 | 33.3 |
| Other | 10 | 16.4 | 6 | 13.6 | 6 | 15.8 | 3 | 12.5 |
| **Inclusion criteria** |  |  |  |  |  |  |  |  |
| Smoking cessation advice | 61 | 100 | n/a | n/a | 38 | 100 | n/a | n/a |
| Two articles or book chapters in the last 5 years | n/a | n/a | 38 | 86.4 | n/a | n/a | 24 | 100 |
| Grant received in the last 5 years | n/a | n/a | 32 | 72.7 | n/a | n/a | 18 | 75 |
| Intervention developed in the last 5 years | n/a | n/a | 32 | 72.7 | n/a | n/a | 21 | 87.5 |
| Five years of experience | n/a | n/a | 31 | 70.5 | n/a | n/a | 19 | 79.2 |
| **Area of expertise** |  |  |  |  |  |  |  |  |
| Experience with DA development | 15 | 24.6 | 19 | 43.2 | 8 | 21.1 | 12 | 50 |
| (Development of) non-digital behavioural interventions for smoking cessation | n/a | n/a | 10 | 22.7 | n/a | n/a | 9 | 37.5 |
| (Development of) digital behavioural interventions for smoking cessation | n/a | n/a | 13 | 29.5 | n/a | n/a | 7 | 29.2 |
| (Development of) pharmaceutical interventions for smoking cessation | n/a | n/a | 5 | 11.4 | n/a | n/a | 3 | 12.5 |
| Determinants of smoking behaviour and tobacco use in general | n/a | n/a | 9 | 20.5 | n/a | n/a | 4 | 16.7 |
| Other | n/a | n/a | 7 | 15.9 | n/a | n/a | 1 | 4.2 |

**Note.** Percentage always refer to the respective samples (i.e., smoking cessation counsellors or scientific experts); n/a = questions were not posed to this group

## References

Abhyankar, P., Volk, R. J., Blumenthal-Barby, J., Bravo, P., Buchholz, A., Ozanne, E., Vidal, D. C., Col, N., & Stalmeier, P. (2013). Balancing the presentation of information and options in patient decision aids: An updated review. BMC Medical Informatics and Decision Making, 13(2), S6. https://doi.org/10.1186/1472-6947-13-s2-s6

Ajzen, I. (1991). The theory of planned behavior. Organizational Behavior and Human Decision Processes, 50(2), 179–211. https://doi.org/10.1016/0749-5978(91)90020-T

Bansal, M. A., Cummings, K. M., Hyland, A., & Giovino, G. A. (2004). Stop-smoking medications: Who uses them, who misuses them, and who is misinformed about them? Nicotine & Tobacco Research, 6. https://doi.org/10.1080/14622200412331320707

Bekker, H., Thornton, J. G., Airey, C. M., Connelly, J. B., Hewison, J., Robinson, M. B., Lilleyman, J., MacIntosh, M., Maule, A. J., & Michie, S. (1999). Informed decision making: An annotated bibliography and systematic review. Health Technol Assess, 3(1), 1–156.

BinDhim, N. F., McGeechan, K., & Trevena, L. (2018). Smartphone Smoking Cessation Application (SSC App) trial: A multicountry double-blind automated randomised controlled trial of a smoking cessation decision-aid ‘app’. BMJ Open, 8(1), e017105. https://doi.org/10.1136/bmjopen-2017-017105

Bol, N., Høie, N. M., Nguyen, M. H., & Smit, E. S. (2019). Customization in mobile health apps: Explaining effects on physical activity intentions by the need for autonomy. Digital Health, 5, 2055207619888074. https://doi.org/10.1177/2055207619888074

Borland, R., Li, L., Driezen, P., Wilson, N., Hammond, D., Thompson, M. E., Fong, G. T., Mons, U., Willemsen, M. C., McNeill, A., Thrasher, J. F., & Cummings, K. M. (2012). Cessation assistance reported by smokers in 15 countries participating in the International Tobacco Control (ITC) policy evaluation surveys. Addiction, 107(1), 197–205. https://doi.org/10.1111/j.1360-0443.2011.03636.x

Brazier, J. E., Dixon, S., & Ratcliffe, J. (2009). The role of patient preferences in cost-effectiveness analysis. Pharmacoeconomics, 27(9), 705. https://doi.org/10.2165/11314840-000000000-00000

Cahill, K., Stevens, S., Perera, R., & Lancaster, T. (2013). Pharmacological interventions for smoking cessation: An overview and network meta-analysis. Cochrane Database of Systematic Reviews, 5. https://doi.org/10.1002/14651858.CD009329.pub2

Chavannes, N., Drenthen, T., Wind, L., Van Avendonk, M., Van den Donk, M., & Verduijn, M. (2017). NHG-Behandelrichtlijn Stoppen met roken.

Cokkinides, V. E., Ward, E., Jemal, A., & Thun, M. J. (2005). Under-use of smoking-cessation treatments: Results from the National Health Interview Survey, 2000. American Journal of Preventive Medicine, 28(1), 119–122. https://doi.org/10.1016/j.amepre.2004.09.007

Coulter, A., Stilwell, D., Kryworuchko, J., Mullen, P. D., Ng, C. J., & van der Weijden, T. (2013). A systematic development process for patient decision aids. BMC Medical Informatics and Decision Making, 13(2), S2. https://doi.org/10.1186/1472-6947-13-S2-S2

Cupertino, A. P., Richter, K., Cox, L. S., Garrett, S., Ramirez, R., Mujica, F., & Ellerbeck, E. F. (2010). Feasibility of a Spanish/English Computerized Decision Aid to Facilitate Smoking Cessation Efforts in Underserved Communities. Journal of Health Care for the Poor and Underserved, 21(2), 504–517. https://doi.org/10.1353/hpu.0.0307

Davis, S., Huebner, A., Piercy, F., Shettler, L., Meszaros, P. S., & Matheson, J. (2004). Female Adolescent Smoking: A Delphi Study on Best Prevention Practices. Journal of Drug Education, 34(3), 295–311. https://doi.org/10.2190/M8C4-HF1G-153K-TM6E

de Ruijter, D., Smit, E. S., de Vries, H., Goossens, L., & Hoving, C. (2017). Understanding Dutch practice nurses’ adherence to evidence-based smoking cessation guidelines and their needs for web-based adherence support: Results from semistructured interviews. BMJ Open, 7(3), e014154. https://doi.org/10.1136/bmjopen-2016-014154

de Vries, H., & Brug, J. (1999). Computer-tailored interventions motivating people to adopt health promoting behaviours: Introduction to a new approach. Patient Education and Counseling, 36, 99–105. https://doi.org/10.1016/S0738-3991(98)00127-X

DeMeester, R. H., Lopez, F. Y., Moore, J. E., Cook, S. C., & Chin, M. H. (2016). A Model of Organizational Context and Shared Decision Making: Application to LGBT Racial and Ethnic Minority Patients. Journal of General Internal Medicine, 31(6), 651–662. https://doi.org/10.1007/s11606-016-3608-3

Durand, M. A., Witt, J., Joseph-Williams, N., Newcombe, R. G., Politi, M. C., Sivell, S., & Elwyn, G. (2015). Minimum standards for the certification of patient decision support interventions: Feasibility and application. Patient Education and Counseling, 98(4), 462–468. https://doi.org/10.1016/j.pec.2014.12.009

Fagerlin, A., Pignone, M., Abhyankar, P., Col, N., Feldman-Stewart, D., Gavaruzzi, T., Kryworuchko, J., Levin, C. A., Pieterse, A. H., Reyna, V., Stiggelbout, A., Scherer, L. D., Wills, C., & Witteman, H. O. (2013). Clarifying values: An updated review. BMC Medical Informatics and Decision Making, 13(2), S8. https://doi.org/10.1186/1472-6947-13-S2-S8

Ferreira, P. L., Tavares, A. I., Quintal, C., & Santana, P. (2018). EU health systems classification: A new proposal from EURO-HEALTHY. BMC Health Services Research, 18(1), 511. https://doi.org/10.1186/s12913-018-3323-3

Flycatcher Internet Research. (2018). Home—Flycatcher. https://www.flycatcher.eu/nl/

Freund, T., Everett, C., Griffiths, P., Hudon, C., Naccarella, L., & Laurant, M. (2015). Skill mix, roles and remuneration in the primary care workforce: Who are the healthcare professionals in the primary care teams across the world? International Journal of Nursing Studies, 52(3), 727– 743. https://doi.org/10.1016/j.ijnurstu.2014.11.014

Gale, N. K., Heath, G., Cameron, E., Rashid, S., & Redwood, S. (2013). Using the framework method for the analysis of qualitative data in multi-disciplinary health research. BMC Medical Research Methodology, 13(1), 117. https://doi.org/10.1186/1471-2288-13-117

Galesic, M., Garcia-Retamero, R., & Gigerenzer, G. (2009). Using icon arrays to communicate medical risks: Overcoming low numeracy. Health Psychology, 28(2), 210–216. https://doi.org/10.1037/a0014474

Genootschap, N. H., & Vereniging, L. H. (2011). NHG/LHV Standpunt: Het (ondersteunend) Team in de huisartsenvoorziening. Utrecht: Nederlands Huisartsen Genootschap and Landelijke Huisartsen Vereniging.

Gültzow, T., Smit, E. S., Hudales, R., Dirksen, C. D., & Hoving, C. (2020). Smoker profiles and their influence on smokers’ intention to use a digital decision aid aimed at the uptake of evidence-based smoking cessation tools: An explorative study. DIGITAL HEALTH, 6, 2055207620980241. https://doi.org/10.1177/2055207620980241

Gültzow, T., Smit, E. S., Hudales, R., Knapen, V., Rademakers, J., Dirksen, C. D., & Hoving, C. (2020). An Autonomy-Supportive Online Decision Aid to Assist Smokers in Choosing Evidence-Based Cessation Assistance: Development Process and Protocol of a Randomized Controlled Trial. JMIR Res Protoc, 9(12), e21772. https://doi.org/10.2196/21772

Gültzow, T., Zijlstra, D., Bolman, C., de Vries, H., Dirksen, C. D., Muris, J. W. M., Smit, E. S., & Hoving, C. (2021). Decision aids to facilitate decision making around behavior change in the field of health promotion: A scoping review. Patient Education and Counseling. https://doi.org/10.1016/j.pec.2021.01.015

Hoffman, A. S., Volk, R. J., Saarimaki, A., Stirling, C., Li, L. C., Härter, M., Kamath, G. R., & Llewellyn-Thomas, H. (2013). Delivering patient decision aids on the Internet: Definitions, theories, current evidence, and emerging research areas. BMC Medical Informatics and Decision Making, 13(2), S13. https://doi.org/10.1186/1472-6947-13-s2-s13

Hooiveld, T., Molenaar, J. M., van der Heijde, C. M., Meijman, F. J., Groen, T. P., & Vonk, P. (2018). End-user involvement in developing and field testing an online contraceptive decision aid. SAGE Open Medicine, 6, 2050312118809462. https://doi.org/10.1177/2050312118809462

IBM Corp. (2017). BM SPSS Statistics for Macintosh, Version 25.0. IBM Corp.

Kahende, J. W., Loomis, B. R., Adhikari, B., & Marshall, L. (2009). A Review of Economic Evaluations of Tobacco Control Programs. International Journal of Environmental Research and Public Health, 6(1). https://doi.org/10.3390/ijerph6010051

Mason, M. (2010). Sample Size and Saturation in PhD Studies Using Qualitative Interviews. FQS, 11(3). https://doi.org/10.17169/fqs-11.3.1428

Matkin, W., Ordóñez-Mena, J. M., & Hartmann-Boyce, J. (2019). Telephone counselling for smoking cessation. Cochrane Database of Systematic Reviews, 5. https://doi.org/10.1002/14651858.CD002850.pub4

Moyo, F., Archibald, E., & Slyer, J. T. (2018). Effectiveness of decision aids for smoking cessation in adults: A quantitative systematic review. JBI Evidence Synthesis, 16(9). https://doi.org/10.11124/JBISRIR-2017-003698

QSR International. (2018). NVivo (Version 12) [MacOS].

Rice, V. H., Heath, L., Livingstone-Banks, J., & Hartmann-Boyce, J. (2017). Nursing interventions for smoking cessation. Cochrane Database of Systematic Reviews, 12. https://doi.org/10.1002/14651858.CD001188.pub5

Risk Science Center and Center for Bioethics and Social Sciences in Medicine, University of Michigan. (n.d.). Iconarray.com. Retrieved January 1, 2020, from http://www.iconarray.com/

Ruger, J. P., & Lazar, C. M. (2012). Economic Evaluation of Pharmaco-and Behavioral Therapies for Smoking Cessation: A Critical and Systematic Review of Empirical Research. Annual Review of Public Health, 33, 279–305. https://doi.org/10.1146/annurev-publhealth-031811-124553

Ryan, R. M., & Deci, E. L. (2008). Self-determination theory and the role of basic psychological needs in personality and the organization of behavior. In Handbook of personality: Theory and research, 3rd ed. (pp. 654–678). The Guilford Press.

Samet, J. M. (2013). Tobacco smoking: The leading cause of preventable disease worldwide. Thoracic Surgery Clinics, 23(2), 103–112. https://doi.org/10.1016/j.thorsurg.2013.01.009

Schneider, F., van Osch, L., & de Vries, H. (2012). Identifying Factors for Optimal Development of Health-Related Websites: A Delphi Study Among Experts and Potential Future Users. Journal of Medical Internet Research, 14(1), e18. PMC. https://doi.org/10.2196/jmir.1863

Scott, S. G., & Bruce, R. A. (1995). Decision-Making Style: The Development and Assessment of a New Measure. Educational and Psychological Measurement, 55(5), 818–831. https://doi.org/10.1177/0013164495055005017

Stacey, D., Légaré, F., Lewis, K., Barry, M. J., Bennett, C. L., Eden, K. B., Holmes-Rovner, M., Llewellyn-Thomas, H., Lyddiatt, A., Thomson, R., & Trevena, L. (2017). Decision aids for people facing health treatment or screening decisions. Cochrane Database of Systematic Reviews, 4. https://doi.org/10.1002/14651858.CD001431.pub5

Stead, L. F., Buitrago, D., Preciado, N., Sanchez, G., Hartmann-Boyce, J., & Lancaster, T. (2013). Physician advice for smoking cessation. Cochrane Database of Systematic Reviews, 5, CD000165. https://doi.org/10.1002/14651858.CD000165.pub4

Stead, L. F., Perera, R., Bullen, C., Mant, D., Hartmann-Boyce, J., Cahill, K., & Lancaster, T. (2012). Nicotine replacement therapy for smoking cessation. Cochrane Database of Systematic Reviews, 11, CD000146. https://doi.org/10.1002/14651858.CD000146.pub4

Syrowatka, A., Krömker, D., Meguerditchian, A. N., & Tamblyn, R. (2016). Features of Computer-Based Decision Aids: Systematic Review, Thematic Synthesis, and Meta-Analyses. Journal of Medical Internet Research, 18(1), e20. https://doi.org/10.2196/jmir.4982

Taylor, G. M. J., Dalili, M. N., Semwal, M., Civljak, M., Sheikh, A., & Car, J. (2017). Internet-based interventions for smoking cessation. Cochrane Database of Systematic Reviews, 9. https://doi.org/10.1002/14651858.CD007078.pub5

Vaisson, G., Provencher, T., Dugas, M., Trottier, M. È., Chipenda Dansokho, S., Colquhoun, H., Fagerlin, A., Giguere, A. M. C., Hakim, H., Haslett, L., Hoffman, A. S., Ivers, N. M., Julien, A. S., Légaré, F., Renaud, J. S., Stacey, D., Volk, R. J., & Witteman, H. O. (2021). User Involvement in the Design and Development of Patient Decision Aids and Other Personal Health Tools: A Systematic Review. Medical Decision Making, 41(3), 261–274. https://doi.org/10.1177/0272989X20984134

van den Berg, M., Timmermans, D. R. M., ten Kate, L. P., van Vugt, J. M. G., & van der Wal, G. (2006). Informed decision making in the context of prenatal screening. Patient Education and Counseling, 63(1), 110–117. https://doi.org/10.1016/j.pec.2005.09.007

Wangberg, S. C., Nilsen, O., Antypas, K., & Gram, I. T. (2011). Effect of Tailoring in an Internet-Based Intervention for Smoking Cessation: Randomized Controlled Trial. Journal of Medical Internet Research, 13(4), e121. https://doi.org/10.2196/jmir.1605

Willemsen, M. C., Wiebing, M., Van Emst, A., & Zeeman, G. (2006). Helping smokers to decide on the use of efficacious smoking cessation methods: A randomized controlled trial of a decision aid. Addiction, 101(3), 441–449. https://doi.org/10.1111/j.1360-0443.2006.01349.x

Witteman, H. O., Julien, A. S., Ndjaboue, R., Exe, N. L., Kahn, V. C., Fagerlin, A., & Zikmund-Fisher, B. J. (2020). What Helps People Make Values-Congruent Medical Decisions? Eleven Strategies Tested across 6 Studies. Medical Decision Making, 40(3), 266–278. https://doi.org/10.1177/0272989X20904955

Zhu, S. H., Melcer, T., Sun, J., Rosbrook, B., & Pierce, J. P. (2000). Smoking cessation with and without assistance: A population-based analysis. American Journal of Preventive Medicine, 18(4), 305–311. https://doi.org/10.1016/S0749-3797(00)00124-0

